# A step-by-step Researcher’s Guide to the use of an AI-based transformer in epidemiology: an exploratory analysis of ChatGPT using the STROBE checklist for observational studies

**DOI:** 10.1101/2023.02.06.23285514

**Authors:** Francesco Sanmarchi, Andrea Bucci, Davide Golinelli

**Affiliations:** Department of Biomedical and Neuromotor Sciences, Alma Mater Studiorum – University of Bologna, Bologna, Italy; University of Macerata, Macerata, Italy

**Author notes:** **Corresponding author** Francesco Sanmarchi, MD, Alma Mater Studiorum - Università di Bologna, Via San Giacomo 12, 40126, Bologna, Italy.

**Keywords:** STROBE, ChatGPT, Methodology, Scientific research, Epidemiology, Public Health, AI, Transformers, Ethics, Legal

## Abstract

**Objectives:** This study aims at investigating how early-stage AI-based transformers can support researchers in designing and conducting an epidemiological study. To accomplish this, we used ChatGPT to reformulate the STROBE recommendations into a list of questions to be answered by the transformer itself. We then qualitatively evaluated the coherence and relevance of the transformer’s outputs.

**Study design:** Descriptive study.

**Methods:** We first chose a study to be used as a basis for the simulation. We then used ChatGPT to transform each STROBE checklist’s item into specific prompts. Each answer to the respective prompt was evaluated by independent researchers in terms of coherence and relevance.

**Results:** The mean scores assigned to each prompt were heterogeneous. On average, for the coherence domain, the overall mean score was 3.6 out of 5.0, and for relevance it was 3.3 out of 5.0. The lowest scores were assigned to items belonging to the Methods section of the checklist.

**Conclusions:** ChatGPT can be considered as a valuable support for researchers in conducting an epidemiological study, following internationally recognized guidelines and standards. It is crucial for the users to have knowledge on the subject and a critical mindset when evaluating the outputs. The potential benefits of AI in scientific research and publishing are undeniable, but it is crucial to address the risks, and the ethical and legal consequences associated with its use.

## 1. Introduction

The scientific method is globally recognized as a systematic and logical approach to discovering new knowledge and understanding the natural world, and is also the foundation of all scientific inquiry which is critical for the advancement of human knowledge [1]. It involves posing a hypothesis, collecting data through observation and experimentation, and analyzing and interpreting the results to reach a conclusion. Hence it helps to ensure that research is objective, replicable, and based on empirical evidence. In the field of epidemiology, the scientific method is essential for conducting rigorous and transparent studies that can accurately address public health issues. As in many types of other research, in a typical epidemiologic study, the general flowchart of the scientific method process [2] includes identifying a research question, conducting a literature review, designing the study, collecting and analyzing data, and interpreting the results and implications.

Therefore, given the importance and complexity of this process, it appears essential to define shared rules, standards, and methodologies to guide the conduct of studies and mitigate the risk of misconduct or errors. In other words, it is crucial that epidemiological studies are conducted transparently and rigorously, following acknowledged guidelines. The importance of this has been underlined by many authors [3–5] and led to the development of the most used guidelines globally: the STrengthening the Reporting of OBservational studies in Epidemiology (STROBE) statement and guidelines. The STROBE guidelines provide a framework for the transparent reporting of observational studies, ensuring that the study’s sections (introduction, methods, results, etc.) are accurately and thoroughly reported [2]. Adhering to the STROBE guidelines can increase the reliability and reproducibility of the study, as well as facilitate the critical appraisal and synthesis of the evidence [2].

Artificial intelligence (AI) is a rapidly growing field with numerous applications in a variety of fields [6]. Among these, one where AI has the potential to make a significant impact is clinical research and epidemiology, where it could revolutionize the way these fields operate and significantly improve our understanding and management of public health and healthcare management issues. Both fields rely on the collection and analysis of large amounts of data, making them well-suited for the application of AI technologies [7,8]. One aspect in which AI has yet to show its potential is the overall support for conducting epidemiological studies. In particular, given the complexity of epidemiological studies, and the increasingly present need to speed up and streamline their execution - while preserving scientific quality and integrity - it appears necessary to study applications of AI in supporting the overall conduct of epidemiological studies, i.e. the support for study design, data analysis and interpretation of results. Both the tools and the studies that test and validate them are scarce in this field, and some potentially revolutionary tools, namely AI-based transformers, have only recently been introduced [9–11].

### 1.1. ChatGPT and AI-based transformers

In recent years, techniques based on deep learning are boosting the AI revolution. Among them, Transformers [12,13] show prominent results in a number of applications. A Transformer is a type of neural network architecture that was introduced in the paper “Attention is All You Need” by Vaswani and colleagues in 2017 [14]. The transformer architecture is a type of deep learning model that adopts the mechanism of self-attention and is particularly adept at processing sequential data such as machine translation, language modeling, and text classification. The transformer model incorporates attention mechanisms to weigh the significance of various parts of the input and output sequences, enabling it to effectively capture long-range dependencies and generate more coherent text.

Chat-Generative Pre-training Transformer (ChatGPT) is a recently developed AI model which stands for “controlled hierarchical generative transformer” [15]. It is a machine learning algorithm capable of generating high-quality scientific text [16], that was developed by a team of researchers at OpenAI, and has received significant attention in the scientific community due to its impressive performance and potential applications. AI-based transformers, such as ChatGPT, may represent a significant advance in the field of AI and could have the potential to impact the way scientific research is conducted and communicated. However, there are also several potential drawbacks to their use for scientific research and scientific publishing, particularly in public health and epidemiology, and their use to support researchers in conducting epidemiological studies has not yet been studied.

### 1.2. Aim of the study

The aim of this study is to shed light and describe how early-stage AI-based transformers models can help a researcher in the various steps of an epidemiological study. To do this, we used the OpenAI ChatGPT to partially simulate an epidemiological study, reformulating the STROBE framework into a list of questions to be answered by the transformer itself. We then qualitatively evaluated the coherence and relevance of the transformer’s outputs and discussed the pros and cons of this innovation in the field of epidemiology.

## 2. Methods

In this exploratory study, we assessed how ChatGPT could support an epidemiological study. To do this, we first chose a study to be used as a basis for the simulation, and extracted the research question and the specific aim. We then relied on the STROBE checklist as a basis for formulating the questions to be asked to ChatGPT. Specifically, we asked ChatGPT to transform each STROBE checklist’s item into a question/prompt for itself. We then asked the transformer to answer the questions, in order to provide support to the researcher in conducting the epidemiological study. Each specific answer was evaluated by three independent senior researchers in terms of coherence and relevance for the purposes of conducting the epidemiological study.

When formulating the various prompts, we used the selected study as a guideline [17]. This study by Subramanian and Kumar analyzed the relationship between the percentage of the population fully vaccinated and new cases of COVID-19 in 68 countries. Subramanian study used COVID-19 publicly available data for cross-country analysis. We chose this type of study because its relatively simple design straightforward interpretation.

Starting from this article, we extracted the aim of the study, considering this as a purely human activity related to human curiosity to ask questions and to transform them into research questions. The aim of the study served as the basis for contextualizing the use of AI-based transformers and demonstrating its potential usefulness in supporting the conduct of an epidemiological study.

### 2.1. Using STROBE recommendations prospectively

The STROBE (Strengthening the Reporting of Observational Studies in Epidemiology) guidelines are a set of recommendations for reporting observational studies, including cohort, case-control, and cross-sectional studies. These guidelines provide a framework for transparent and complete reporting of the design, conduct, and results of an observational study, with the aim of improving the quality and transparency of the research and minimizing biases and confounding factors, which can affect the validity and reliability of the results.

The STROBE guidelines can also be used prospectively when conducting an epidemiologic study. By following the STROBE guidelines during the design and conduct of the study, researchers can ensure that the study is conducted in a transparent and rigorous manner and that the results of the study are accurately and clearly reported.

The STROBE guidelines provide specific recommendations for the reporting of various aspects of the study, such as the study design, the sampling and recruitment of participants, the measurement of exposures and outcomes, and the analysis and interpretation of the results.

### 2.3. ChatGPT

GPT-3 (Generative Pre-trained Transformer) is a third-generation, autoregressive language model that uses deep learning to produce human-like text developed by OpenAI. It is designed to generate human-like text and engage in conversation with users naturally and intuitively. To train the GPT model, researchers at OpenAI fed it a large dataset of human-generated text and utilized the model to predict the next word in the sequence. The model was then fine-tuned using backpropagation, a technique that involves adjusting the model’s weights and biases to minimize the error between the predicted output and the actual output. After the model was trained, it was able to generate human-like text by predicting the next word in a sequence based on the context provided by the previous words. The model can be fine-tuned for specific tasks, such as translation or question answering, by training it on a dataset specific to that task.

It is worth noting that ChatGPT, a variant of GPT specifically designed for generating human-like text in a conversational context, such as chatbots or virtual assistants, is not the same as GPT. While ChatGPT is trained on a dataset of human conversation and is able to generate text that is appropriate for use in a chat or messaging context, the underlying technology and principles of how ChatGPT works are the same as those of GPT.

One of the key features of ChatGPT is its ability to generate text that is coherent and follows the structure and style of scientific articles. It has been trained on large datasets by using 175 billion parameters on Microsoft’s Azure’s AI supercomputer and has learned to generate text that is similar to human-written scientific text in terms of grammar, style, and content. This makes it a valuable tool for researchers who need to generate large amounts of scientific text, such as descriptions of experimental methods and results, summaries of research findings, or review articles [18].

### 2.4. The use of ChatGPT for supporting the conduction of the study

First, we identified the purpose of the study as: *[To examine the relationship between the percentage of population fully vaccinated and the incidence of new COVID-19 cases in a given population, and to assess the impact of vaccination on the incidence of COVID-19 cases.]*

To maximize the use of ChatGPT, we adopted the STROBE framework, translating its main items/recommendations into prospective questions for the AI transformer. To standardize the use of the transformer as much as possible, we followed the process listed below:

*STROBE item > question/prompt for ChatGPT > adaptation to the simulated research questions > final answer*

For each STROBE recommendation, we asked the transformer (on December 19-23, 2022) the following question:

“Transform the following recommendation from the STROBE (Strengthening the Reporting of Observational Studies in Epidemiology) guidelines into a question for ChatGPT. Recommendation: [*STROBE recommendation*]”

This provided us Answer#1. Then, the next step has been the specific question:

“[*Answer#1*]+[*The study has the following aim: To examine the relationship between the percentage of population fully vaccinated and the incidence of new COVID-19 cases in a given population, and to assess the impact of vaccination on the incidence of COVID-19 cases*]

### 2.5. Expert assessment

We assessed the outputs of the transformer by using human annotators to rate the quality and relevance of the model’s responses. This has been done through a process called “human evaluation,” in which annotators (NN, FT, AB) were asked to rate the responses produced by the model on a 1 to 5 points Likert scale. The annotators were asked to consider factors such as the coherence of the response and its relevance to the prompt.Given that there are no standardized questions that are specifically designed to evaluate the quality and relevance of the responses produced by ChatGPT or any other language model in terms of coherence and relevance, we evaluated the outputs by asking the expert annotators to consider the following questions:

1. 1.Is the response coherent and well-formed, or is it difficult to understand? (where 5 is the maximum level of coherence)
2. 2.Is the response relevant and informative, or is it lacking in substance? (where 5 is the maximum level of relevance)

For each STROBE recommendation, we formulated a prompt for ChatGPT contextualizing it with the aim of the study by Subramanian and Kumar (2021) [17]. Each response to the prompts was then evaluated by domain experts to assess its consistency and relevance.

## 3. Results

The STROBE checklist includes a list of 35 recommendations (Table S1). From these recommendations we derived 35 specific questions for ChatGPT. Three domain experts evaluated the answers independently using two coherence and relevance scores (Table 1). The mean scores were heterogeneous. On average, for the coherence scores, the overall mean score was 3.6 out of 5.0, and for relevance it was 3.3 out of 5.0.

**Table 1.**
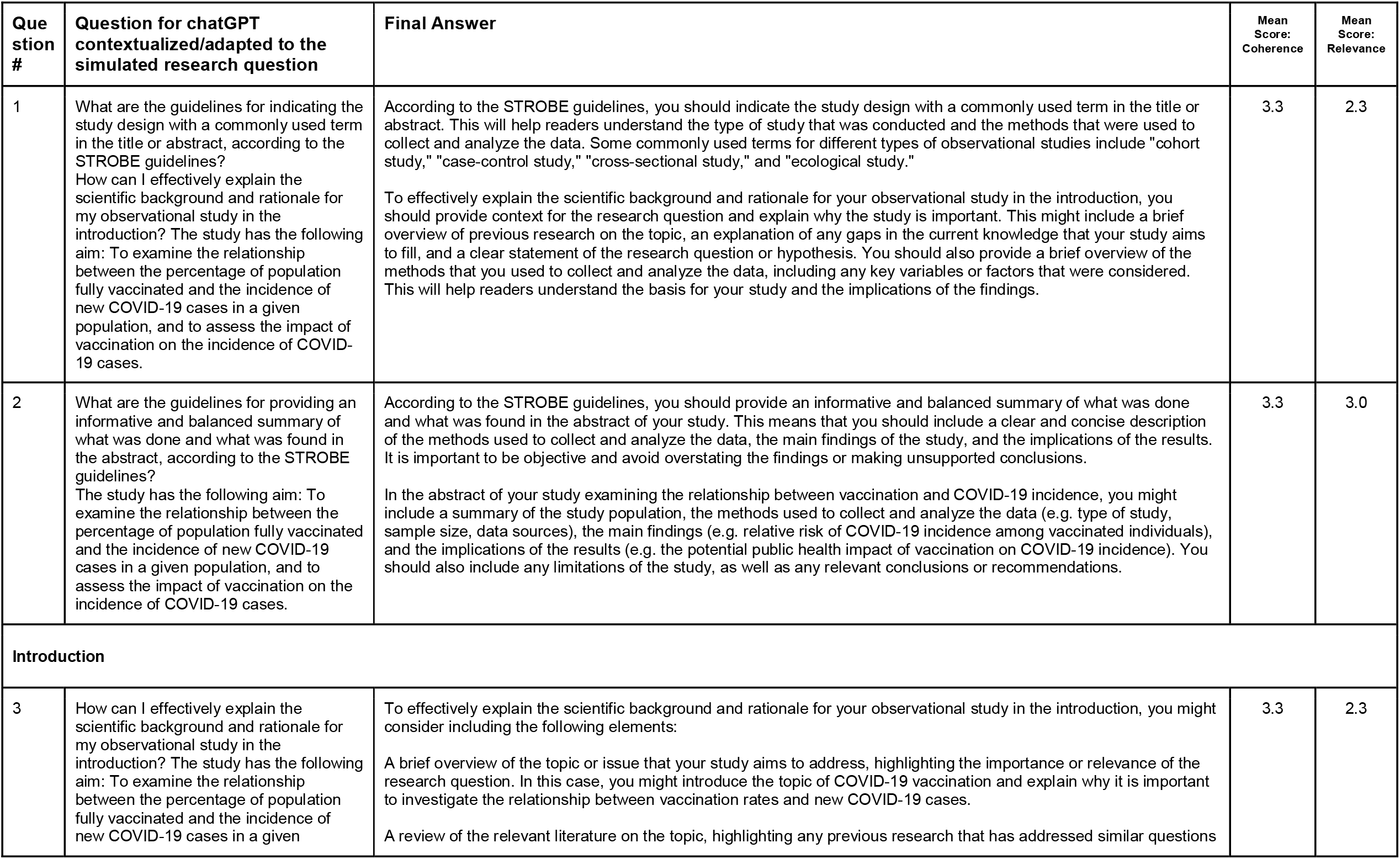
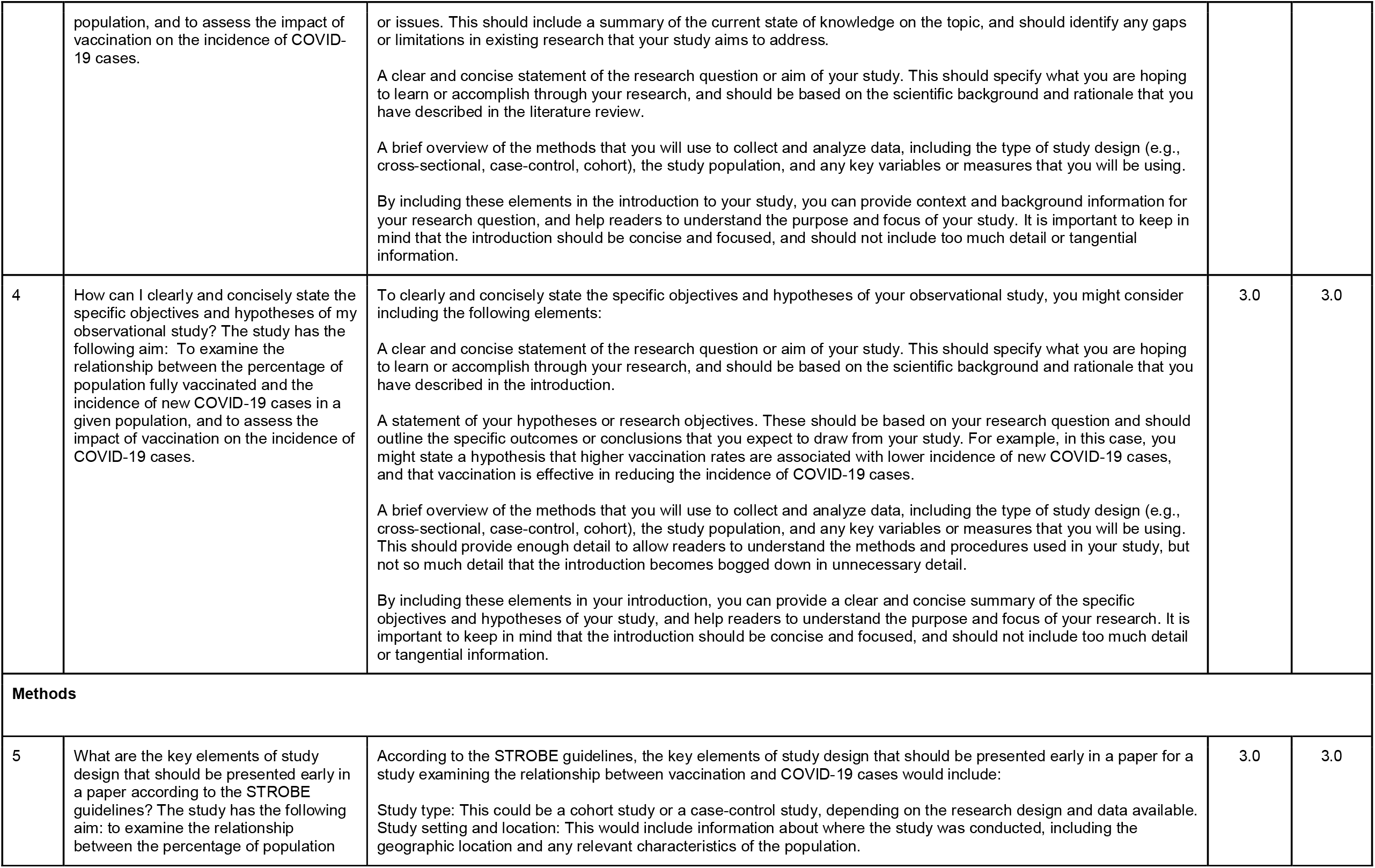
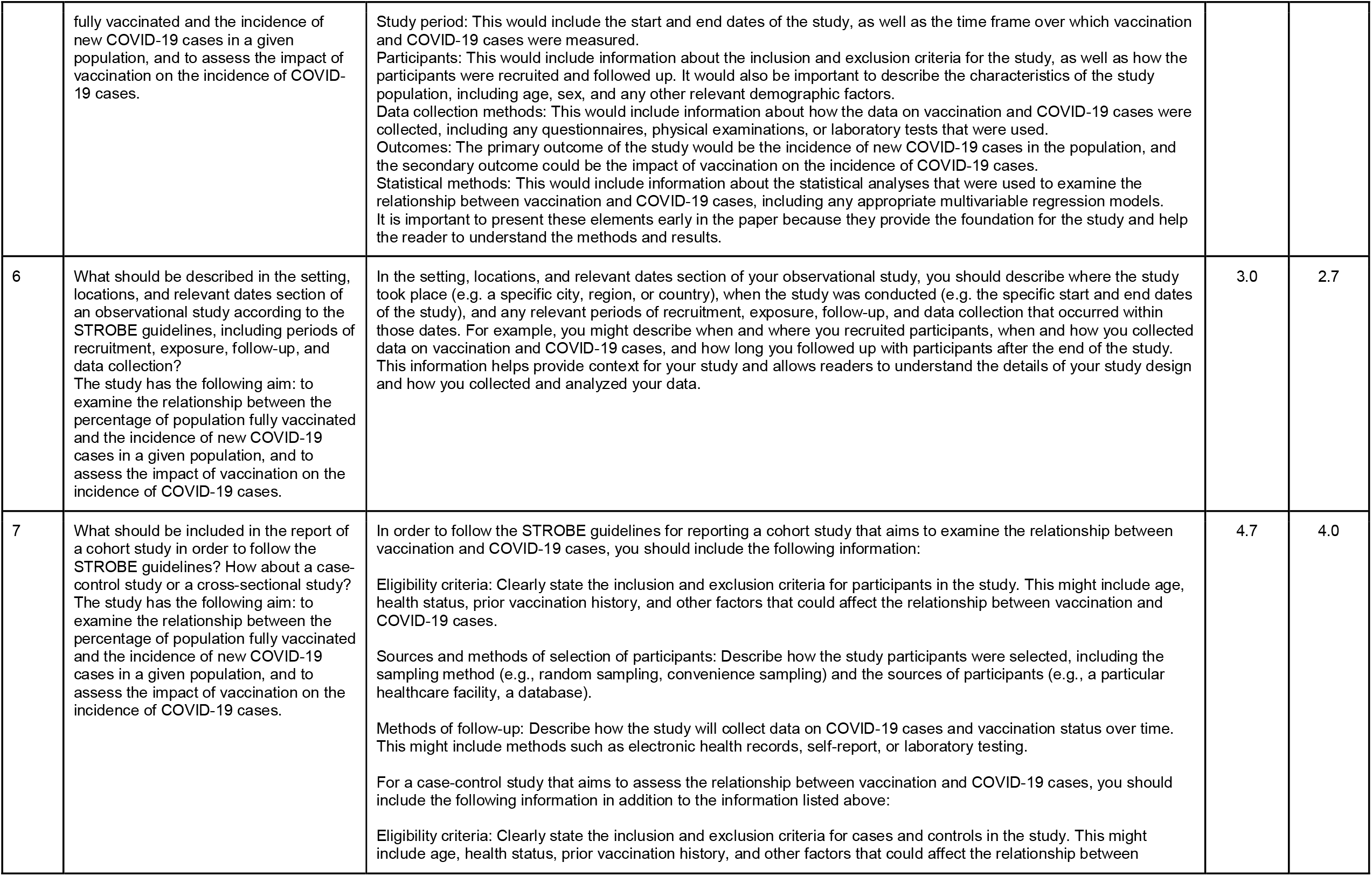
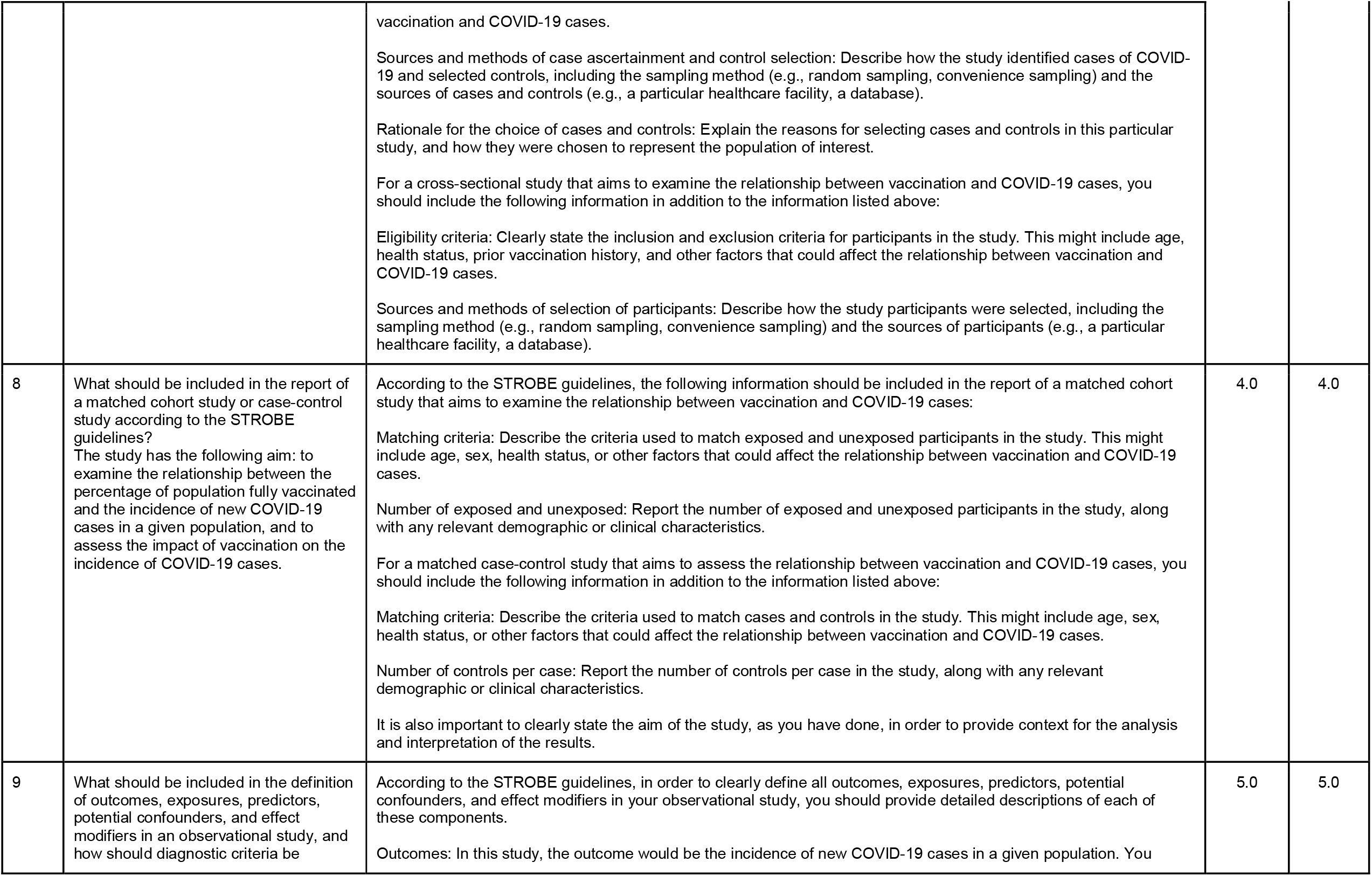
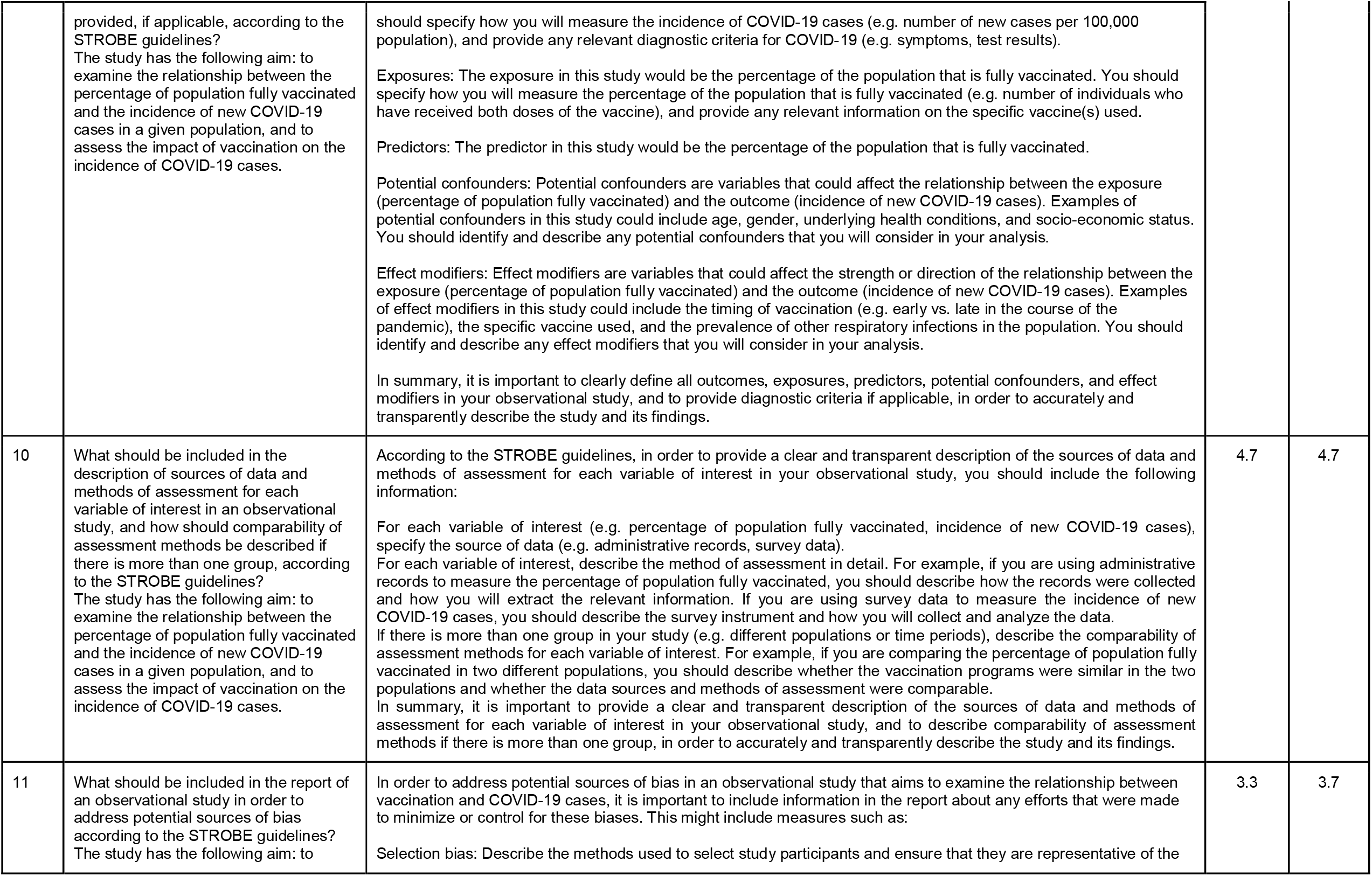
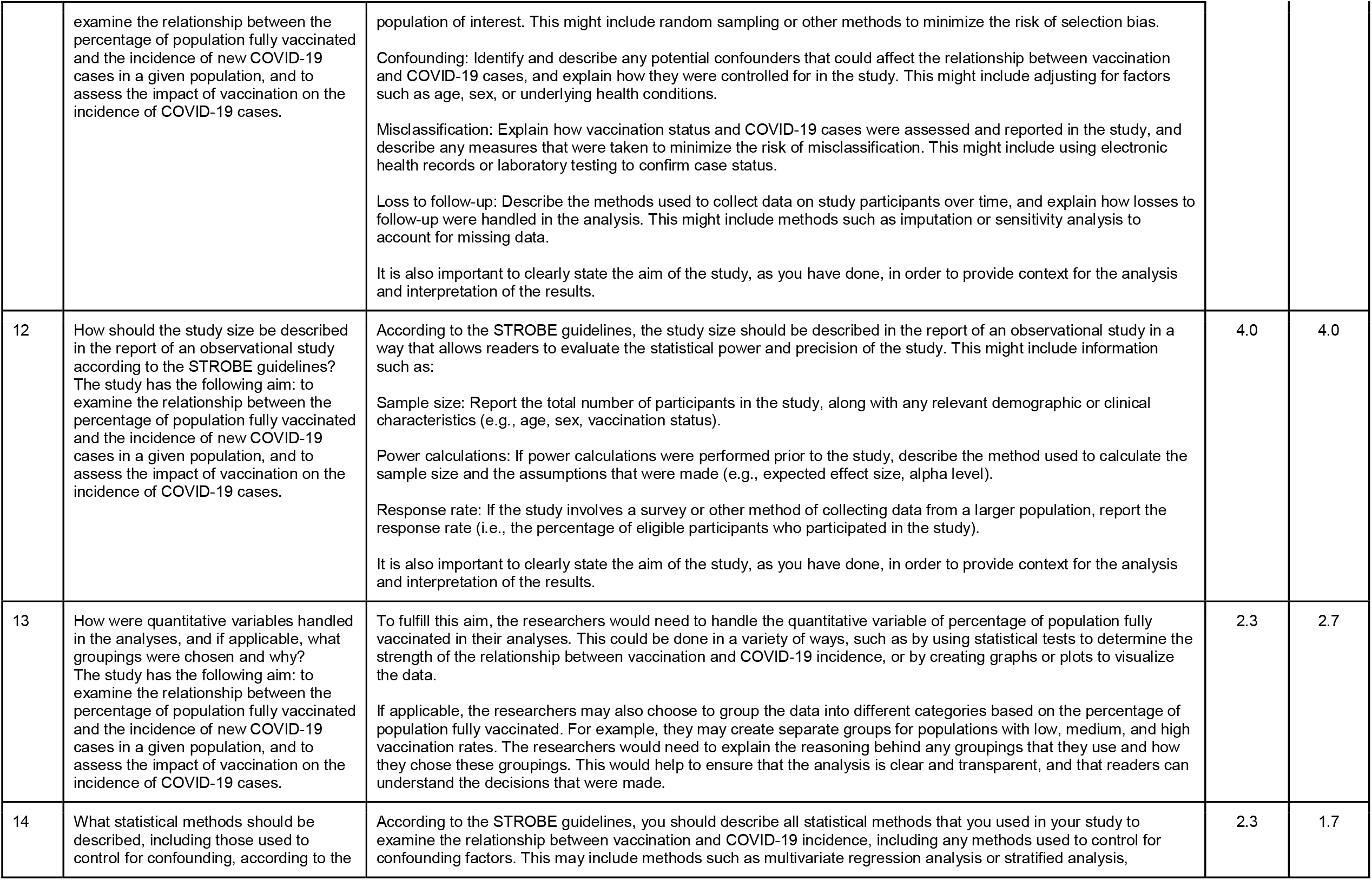
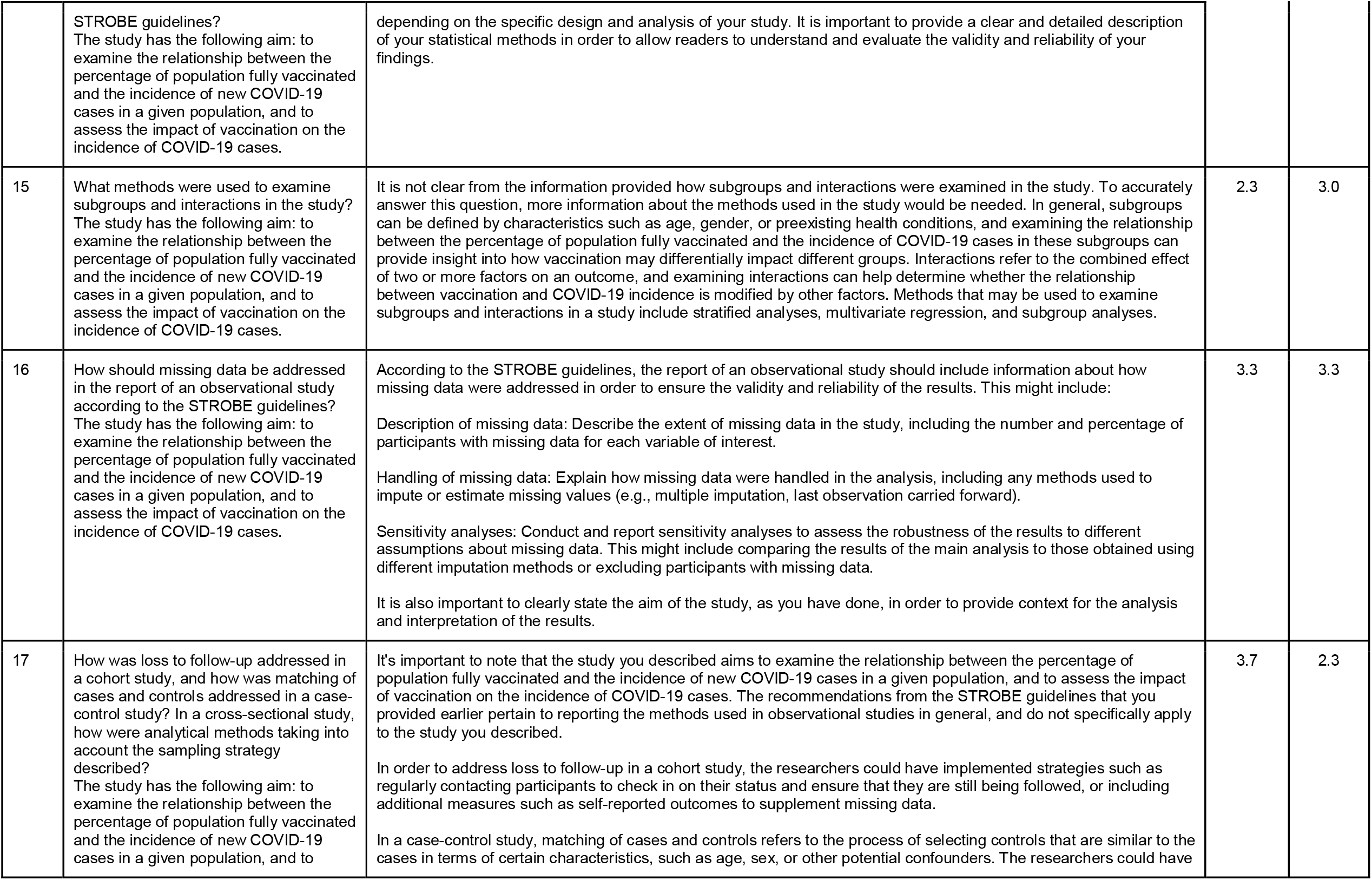
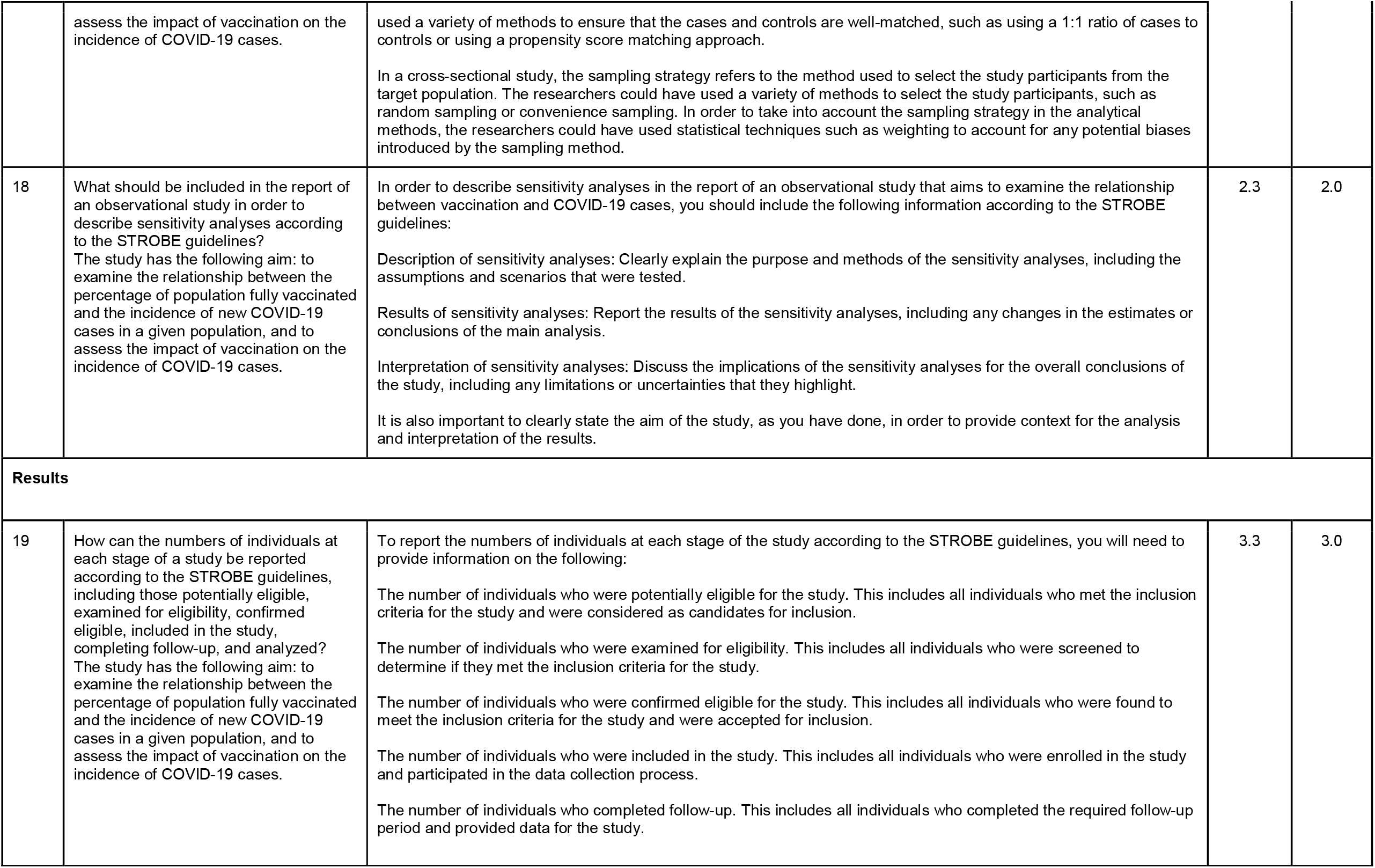
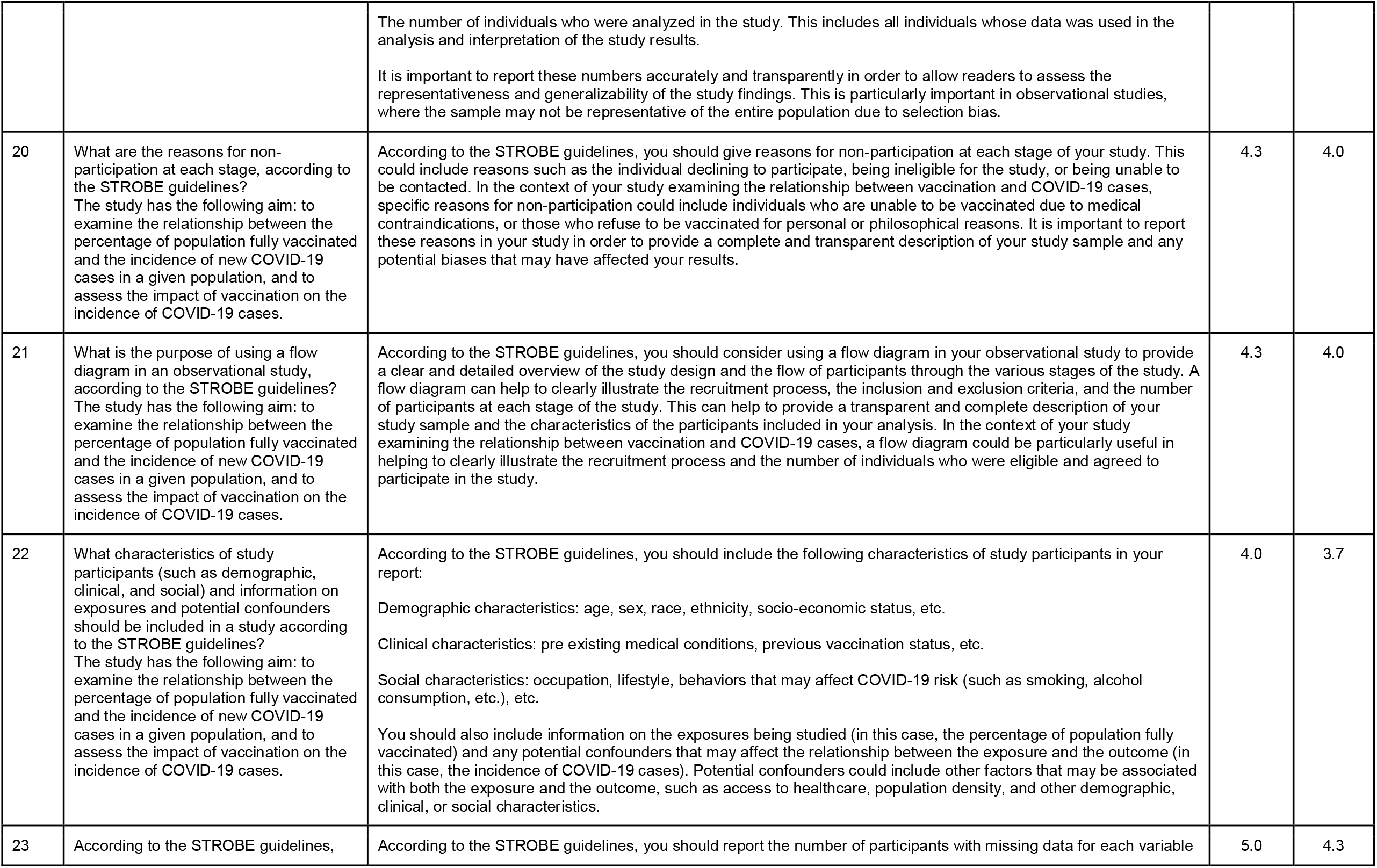
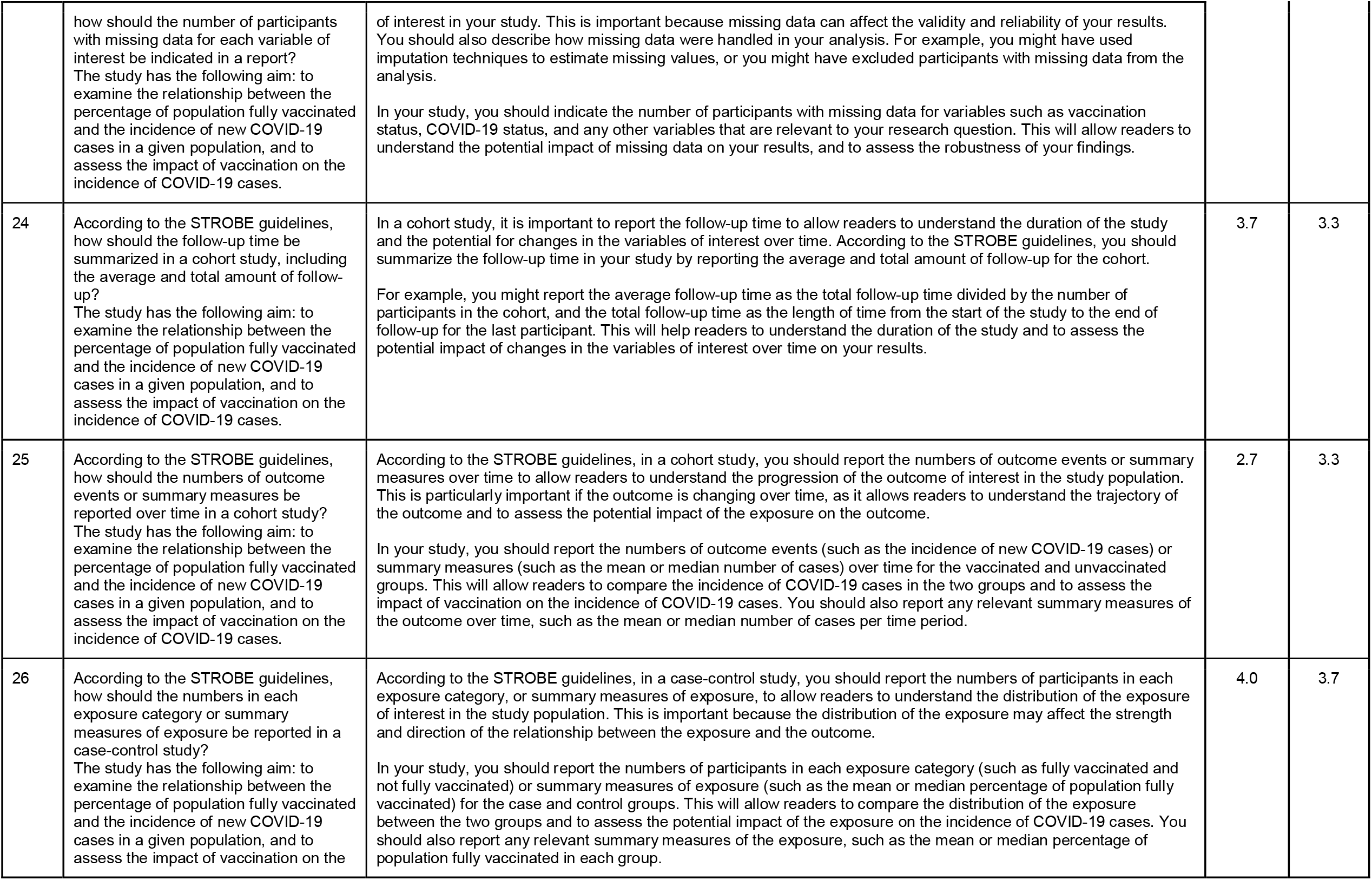
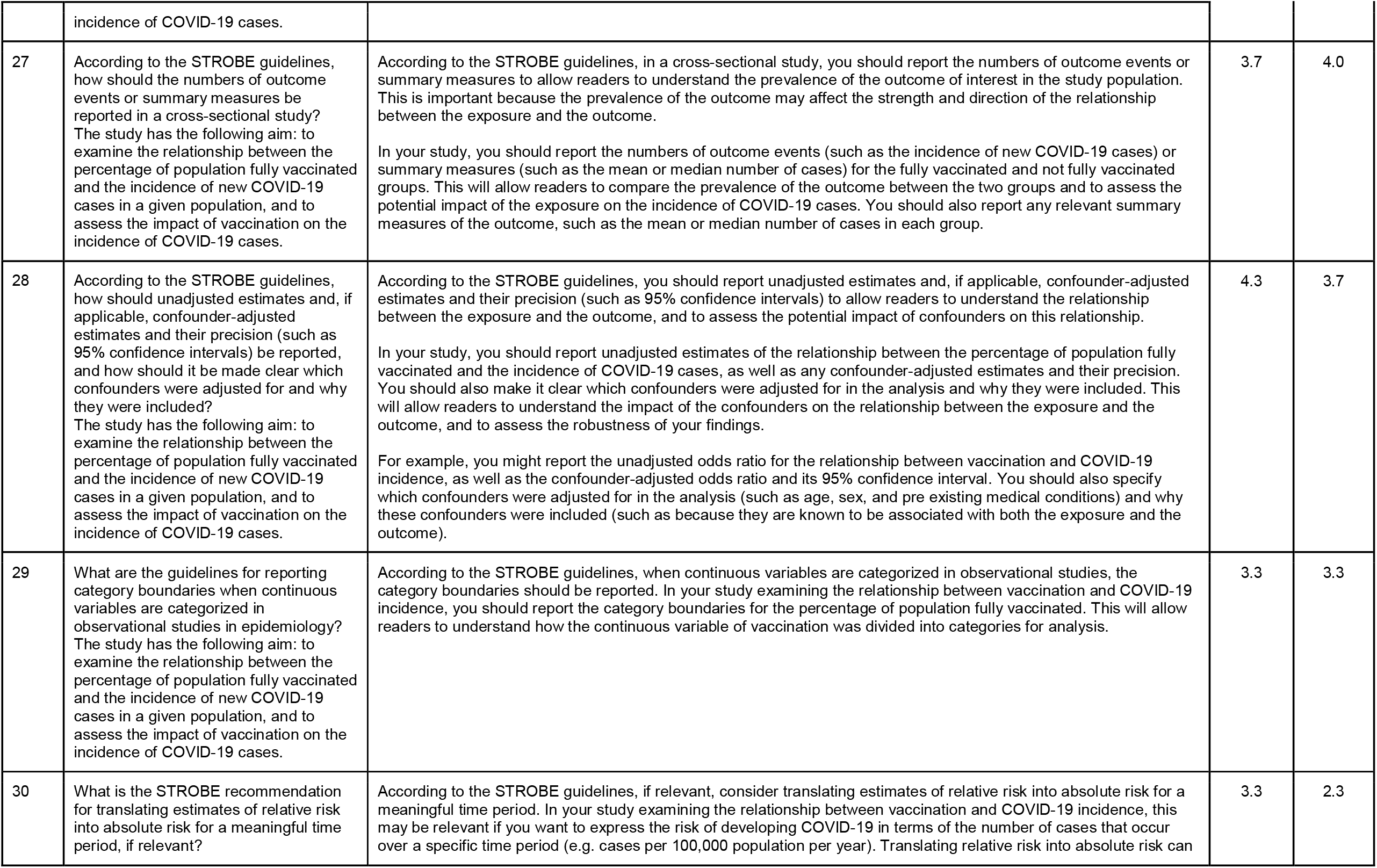
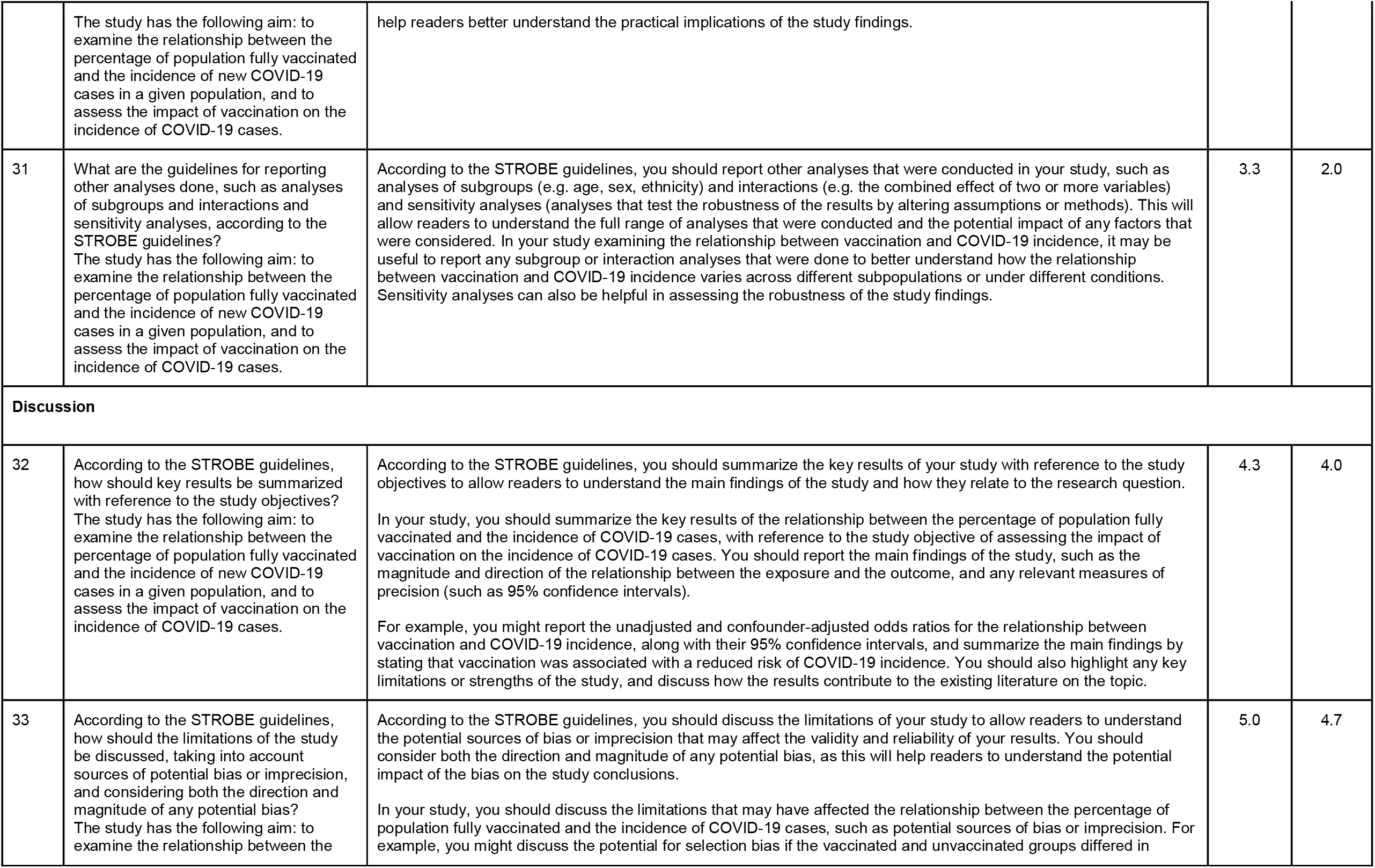
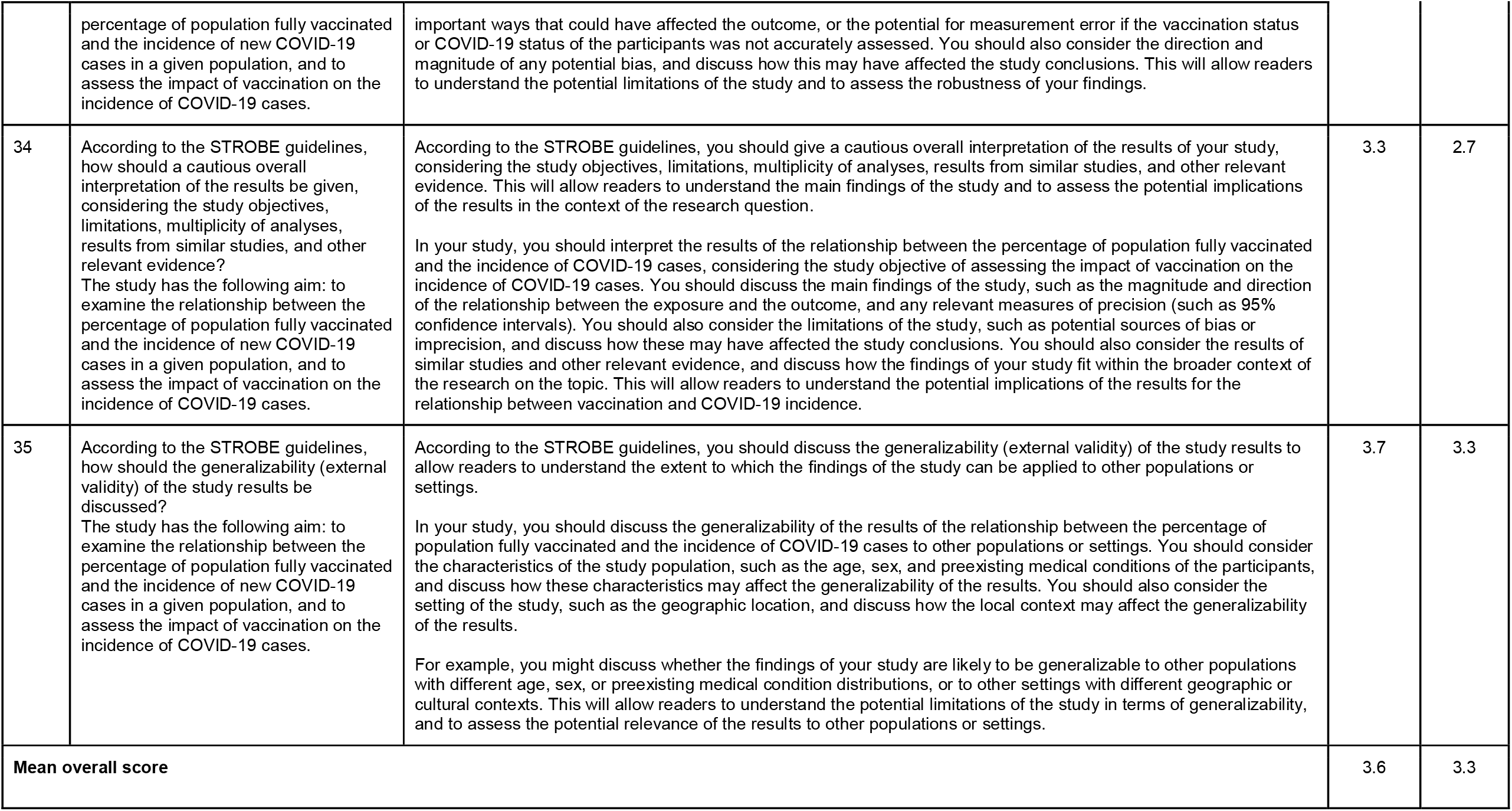
Final Q&A and respective scores.

| **Question #** | **Question for chatGPT contextualized/adapted to the simulated research question** | **Final Answer** | **Mean Score: Coherence** | **Mean Score: Relevance** |
|---|---|---|---|---|
| 1 | What are the guidelines for indicating the study design with a commonly used term in the title or abstract, according to the STROBE guidelines?<br>How can I effectively explain the scientific background and rationale for my observational study in the introduction? The study has the following aim: To examine the relationship between the percentage of population fully vaccinated and the incidence of new COVID-19 cases in a given population, and to assess the impact of vaccination on the incidence of COVID-19 cases. | According to the STROBE guidelines, you should indicate the study design with a commonly used term in the title or abstract. This will help readers understand the type of study that was conducted and the methods that were used to collect and analyze the data. Some commonly used terms for different types of observational studies include "cohort study," "case-control study," "cross-sectional study," and "ecological study."<br>To effectively explain the scientific background and rationale for your observational study in the introduction, you should provide context for the research question and explain why the study is important. This might include a brief overview of previous research on the topic, an explanation of any gaps in the current knowledge that your study aims to fill, and a clear statement of the research question or hypothesis. You should also provide a brief overview of the methods that you used to collect and analyze the data, including any key variables or factors that were considered. This will help readers understand the basis for your study and the implications of the findings. | 3.3 | 2.3 |
| 2 | What are the guidelines for providing an informative and balanced summary of what was done and what was found in the abstract, according to the STROBE guidelines?<br>The study has the following aim: To examine the relationship between the percentage of population fully vaccinated and the incidence of new COVID-19 cases in a given population, and to assess the impact of vaccination on the incidence of COVID-19 cases. | According to the STROBE guidelines, you should provide an informative and balanced summary of what was done and what was found in the abstract of your study. This means that you should include a clear and concise description of the methods used to collect and analyze the data, the main findings of the study, and the implications of the results. It is important to be objective and avoid overstating the findings or making unsupported conclusions.<br>In the abstract of your study examining the relationship between vaccination and COVID-19 incidence, you might include a summary of the study population, the methods used to collect and analyze the data (e.g. type of study, sample size, data sources), the main findings (e.g. relative risk of COVID-19 incidence among vaccinated individuals), and the implications of the results (e.g. the potential public health impact of vaccination on COVID-19 incidence). You should also include any limitations of the study, as well as any relevant conclusions or recommendations. | 3.3 | 3.0 |
| **Introduction** |  |  |  |  |
| 3 | How can I effectively explain the scientific background and rationale for my observational study in the introduction? The study has the following aim: To examine the relationship between the percentage of population fully vaccinated and the incidence of new COVID-19 cases in a given population, and to assess the impact of vaccination on the incidence of COVID-19 cases. | To effectively explain the scientific background and rationale for your observational study in the introduction, you might consider including the following elements:<br>A brief overview of the topic or issue that your study aims to address, highlighting the importance or relevance of the research question. In this case, you might introduce the topic of COVID-19 vaccination and explain why it is important to investigate the relationship between vaccination rates and new COVID-19 cases.<br>A review of the relevant literature on the topic, highlighting any previous research that has addressed similar questions or issues. This should include a summary of the current state of knowledge on the topic, and should identify any gaps or limitations in existing research that your study aims to address.<br>A clear and concise statement of the research question or aim of your study. This should specify what you are hoping to learn or accomplish through your research, and should be based on the scientific background and rationale that you have described in the literature review.<br>A brief overview of the methods that you will use to collect and analyze data, including the type of study design (e.g., cross-sectional, case-control, cohort), the study population, and any key variables or measures that you will be using.<br>By including these elements in the introduction to your study, you can provide context and background information for your research question, and help readers to understand the purpose and focus of your study. It is important to keep in mind that the introduction should be concise and focused, and should not include too much detail or tangential information. | 3.3 | 2.3 |
| 4 | How can I clearly and concisely state the specific objectives and hypotheses of my observational study? The study has the following aim: To examine the relationship between the percentage of population fully vaccinated and the incidence of new COVID-19 cases in a given population, and to assess the impact of vaccination on the incidence of COVID-19 cases. | To clearly and concisely state the specific objectives and hypotheses of your observational study, you might consider including the following elements:<br>A clear and concise statement of the research question or aim of your study. This should specify what you are hoping to learn or accomplish through your research, and should be based on the scientific background and rationale that you have described in the introduction.<br>A statement of your hypotheses or research objectives. These should be based on your research question and should outline the specific outcomes or conclusions that you expect to draw from your study. For example, in this case, you might state a hypothesis that higher vaccination rates are associated with lower incidence of new COVID-19 cases, and that vaccination is effective in reducing the incidence of COVID-19 cases.<br>A brief overview of the methods that you will use to collect and analyze data, including the type of study design (e.g., cross-sectional, case-control, cohort), the study population, and any key variables or measures that you will be using. This should provide enough detail to allow readers to understand the methods and procedures used in your study, but not so much detail that the introduction becomes bogged down in unnecessary detail.<br>By including these elements in your introduction, you can provide a clear and concise summary of the specific objectives and hypotheses of your study, and help readers to understand the purpose and focus of your research. It is important to keep in mind that the introduction should be concise and focused, and should not include too much detail or tangential information. | 3.0 | 3.0 |
| **Methods** |  |  |  |  |
| 5 | What are the key elements of study design that should be presented early in a paper according to the STROBE guidelines? The study has the following aim: to examine the relationship between the percentage of population fully vaccinated and the incidence of new COVID-19 cases in a given population, and to assess the impact of vaccination on the incidence of COVID-19 cases. | According to the STROBE guidelines, the key elements of study design that should be presented early in a paper for a study examining the relationship between vaccination and COVID-19 cases would include:<br>Study type: This could be a cohort study or a case-control study, depending on the research design and data available.<br>Study setting and location: This would include information about where the study was conducted, including the geographic location and any relevant characteristics of the population.<br>Study period: This would include the start and end dates of the study, as well as the time frame over which vaccination and COVID-19 cases were measured.<br>Participants: This would include information about the inclusion and exclusion criteria for the study, as well as how the participants were recruited and followed up. It would also be important to describe the characteristics of the study population, including age, sex, and any other relevant demographic factors.<br>Data collection methods: This would include information about how the data on vaccination and COVID-19 cases were collected, including any questionnaires, physical examinations, or laboratory tests that were used.<br>Outcomes: The primary outcome of the study would be the incidence of new COVID-19 cases in the population, and the secondary outcome could be the impact of vaccination on the incidence of COVID-19 cases.<br>Statistical methods: This would include information about the statistical analyses that were used to examine the relationship between vaccination and COVID-19 cases, including any appropriate multivariable regression models.<br>It is important to present these elements early in the paper because they provide the foundation for the study and help the reader to understand the methods and results. | 3.0 | 3.0 |
| 6 | What should be described in the setting, locations, and relevant dates section of an observational study according to the STROBE guidelines, including periods of recruitment, exposure, follow-up, and data collection?<br>The study has the following aim: to examine the relationship between the percentage of population fully vaccinated and the incidence of new COVID-19 cases in a given population, and to assess the impact of vaccination on the incidence of COVID-19 cases. | In the setting, locations, and relevant dates section of your observational study, you should describe where the study took place (e.g. a specific city, region, or country), when the study was conducted (e.g. the specific start and end dates of the study), and any relevant periods of recruitment, exposure, follow-up, and data collection that occurred within those dates. For example, you might describe when and where you recruited participants, when and how you collected data on vaccination and COVID-19 cases, and how long you followed up with participants after the end of the study. This information helps provide context for your study and allows readers to understand the details of your study design and how you collected and analyzed your data. | 3.0 | 2.7 |
| 7 | What should be included in the report of a cohort study in order to follow the STROBE guidelines? How about a case-control study or a cross-sectional study?<br>The study has the following aim: to examine the relationship between the percentage of population fully vaccinated and the incidence of new COVID-19 cases in a given population, and to assess the impact of vaccination on the incidence of COVID-19 cases. | In order to follow the STROBE guidelines for reporting a cohort study that aims to examine the relationship between vaccination and COVID-19 cases, you should include the following information:<br>Eligibility criteria: Clearly state the inclusion and exclusion criteria for participants in the study. This might include age, health status, prior vaccination history, and other factors that could affect the relationship between vaccination and COVID-19 cases.<br>Sources and methods of selection of participants: Describe how the study participants were selected, including the sampling method (e.g., random sampling, convenience sampling) and the sources of participants (e.g., a particular healthcare facility, a database).<br>Methods of follow-up: Describe how the study will collect data on COVID-19 cases and vaccination status over time. This might include methods such as electronic health records, self-report, or laboratory testing.<br>For a case-control study that aims to assess the relationship between vaccination and COVID-19 cases, you should include the following information in addition to the information listed above:<br>Eligibility criteria: Clearly state the inclusion and exclusion criteria for cases and controls in the study. This might include age, health status, prior vaccination history, and other factors that could affect the relationship between vaccination and COVID-19 cases.<br>Sources and methods of case ascertainment and control selection: Describe how the study identified cases of COVID-19 and selected controls, including the sampling method (e.g., random sampling, convenience sampling) and the sources of cases and controls (e.g., a particular healthcare facility, a database).<br>Rationale for the choice of cases and controls: Explain the reasons for selecting cases and controls in this particular study, and how they were chosen to represent the population of interest.<br>For a cross-sectional study that aims to examine the relationship between vaccination and COVID-19 cases, you should include the following information in addition to the information listed above:<br>Eligibility criteria: Clearly state the inclusion and exclusion criteria for participants in the study. This might include age, health status, prior vaccination history, and other factors that could affect the relationship between vaccination and COVID-19 cases.<br>Sources and methods of selection of participants: Describe how the study participants were selected, including the sampling method (e.g., random sampling, convenience sampling) and the sources of participants (e.g., a particular healthcare facility, a database). | 4.7 | 4.0 |
| 8 | What should be included in the report of a matched cohort study or case-control study according to the STROBE guidelines?<br>The study has the following aim: to examine the relationship between the percentage of population fully vaccinated and the incidence of new COVID-19 cases in a given population, and to assess the impact of vaccination on the incidence of COVID-19 cases. | According to the STROBE guidelines, the following information should be included in the report of a matched cohort study that aims to examine the relationship between vaccination and COVID-19 cases:<br>Matching criteria: Describe the criteria used to match exposed and unexposed participants in the study. This might include age, sex, health status, or other factors that could affect the relationship between vaccination and COVID-19 cases.<br>Number of exposed and unexposed: Report the number of exposed and unexposed participants in the study, along with any relevant demographic or clinical characteristics.<br>For a matched case-control study that aims to assess the relationship between vaccination and COVID-19 cases, you should include the following information in addition to the information listed above:<br>Matching criteria: Describe the criteria used to match cases and controls in the study. This might include age, sex, health status, or other factors that could affect the relationship between vaccination and COVID-19 cases.<br>Number of controls per case: Report the number of controls per case in the study, along with any relevant demographic or clinical characteristics.<br>It is also important to clearly state the aim of the study, as you have done, in order to provide context for the analysis and interpretation of the results. | 4.0 | 4.0 |
| 9 | What should be included in the definition of outcomes, exposures, predictors, potential confounders, and effect modifiers in an observational study, and how should diagnostic criteria be provided, if applicable, according to the STROBE guidelines?<br>The study has the following aim: to examine the relationship between the percentage of population fully vaccinated and the incidence of new COVID-19 cases in a given population, and to assess the impact of vaccination on the incidence of COVID-19 cases. | According to the STROBE guidelines, in order to clearly define all outcomes, exposures, predictors, potential confounders, and effect modifiers in your observational study, you should provide detailed descriptions of each of these components.<br>Outcomes: In this study, the outcome would be the incidence of new COVID-19 cases in a given population. You should specify how you will measure the incidence of COVID-19 cases (e.g. number of new cases per 100,000 population), and provide any relevant diagnostic criteria for COVID-19 (e.g. symptoms, test results).<br>Exposures: The exposure in this study would be the percentage of the population that is fully vaccinated. You should specify how you will measure the percentage of the population that is fully vaccinated (e.g. number of individuals who have received both doses of the vaccine), and provide any relevant information on the specific vaccine(s) used.<br>Predictors: The predictor in this study would be the percentage of the population that is fully vaccinated.<br>Potential confounders: Potential confounders are variables that could affect the relationship between the exposure (percentage of population fully vaccinated) and the outcome (incidence of new COVID-19 cases). Examples of potential confounders in this study could include age, gender, underlying health conditions, and socio-economic status. You should identify and describe any potential confounders that you will consider in your analysis.<br>Effect modifiers: Effect modifiers are variables that could affect the strength or direction of the relationship between the exposure (percentage of population fully vaccinated) and the outcome (incidence of new COVID-19 cases). Examples of effect modifiers in this study could include the timing of vaccination (e.g. early vs. late in the course of the pandemic), the specific vaccine used, and the prevalence of other respiratory infections in the population. You should identify and describe any effect modifiers that you will consider in your analysis.<br>In summary, it is important to clearly define all outcomes, exposures, predictors, potential confounders, and effect modifiers in your observational study, and to provide diagnostic criteria if applicable, in order to accurately and transparently describe the study and its findings. | 5.0 | 5.0 |
| 10 | What should be included in the description of sources of data and methods of assessment for each variable of interest in an observational study, and how should comparability of assessment methods be described if there is more than one group, according to the STROBE guidelines?<br>The study has the following aim: to examine the relationship between the percentage of population fully vaccinated and the incidence of new COVID-19 cases in a given population, and to assess the impact of vaccination on the incidence of COVID-19 cases. | According to the STROBE guidelines, in order to provide a clear and transparent description of the sources of data and methods of assessment for each variable of interest in your observational study, you should include the following information:<br>For each variable of interest (e.g. percentage of population fully vaccinated, incidence of new COVID-19 cases), specify the source of data (e.g. administrative records, survey data).<br>For each variable of interest, describe the method of assessment in detail. For example, if you are using administrative records to measure the percentage of population fully vaccinated, you should describe how the records were collected and how you will extract the relevant information. If you are using survey data to measure the incidence of new COVID-19 cases, you should describe the survey instrument and how you will collect and analyze the data.<br>If there is more than one group in your study (e.g. different populations or time periods), describe the comparability of assessment methods for each variable of interest. For example, if you are comparing the percentage of population fully vaccinated in two different populations, you should describe whether the vaccination programs were similar in the two populations and whether the data sources and methods of assessment were comparable.<br>In summary, it is important to provide a clear and transparent description of the sources of data and methods of assessment for each variable of interest in your observational study, and to describe comparability of assessment methods if there is more than one group, in order to accurately and transparently describe the study and its findings. | 4.7 | 4.7 |
| 11 | What should be included in the report of an observational study in order to address potential sources of bias according to the STROBE guidelines?<br>The study has the following aim: to examine the relationship between the percentage of population fully vaccinated and the incidence of new COVID-19 cases in a given population, and to assess the impact of vaccination on the incidence of COVID-19 cases. | In order to address potential sources of bias in an observational study that aims to examine the relationship between vaccination and COVID-19 cases, it is important to include information in the report about any efforts that were made to minimize or control for these biases. This might include measures such as:<br>Selection bias: Describe the methods used to select study participants and ensure that they are representative of the population of interest. This might include random sampling or other methods to minimize the risk of selection bias.<br>Confounding: Identify and describe any potential confounders that could affect the relationship between vaccination and COVID-19 cases, and explain how they were controlled for in the study. This might include adjusting for factors such as age, sex, or underlying health conditions.<br>Misclassification: Explain how vaccination status and COVID-19 cases were assessed and reported in the study, and describe any measures that were taken to minimize the risk of misclassification. This might include using electronic health records or laboratory testing to confirm case status.<br>Loss to follow-up: Describe the methods used to collect data on study participants over time, and explain how losses to follow-up were handled in the analysis. This might include methods such as imputation or sensitivity analysis to account for missing data.<br>It is also important to clearly state the aim of the study, as you have done, in order to provide context for the analysis and interpretation of the results. | 3.3 | 3.7 |
| 12 | How should the study size be described in the report of an observational study according to the STROBE guidelines?<br>The study has the following aim: to examine the relationship between the percentage of population fully vaccinated and the incidence of new COVID-19 cases in a given population, and to assess the impact of vaccination on the incidence of COVID-19 cases. | According to the STROBE guidelines, the study size should be described in the report of an observational study in a way that allows readers to evaluate the statistical power and precision of the study. This might include information such as:<br>Sample size: Report the total number of participants in the study, along with any relevant demographic or clinical characteristics (e.g., age, sex, vaccination status).<br>Power calculations: If power calculations were performed prior to the study, describe the method used to calculate the sample size and the assumptions that were made (e.g., expected effect size, alpha level).<br>Response rate: If the study involves a survey or other method of collecting data from a larger population, report the response rate (i.e., the percentage of eligible participants who participated in the study).<br>It is also important to clearly state the aim of the study, as you have done, in order to provide context for the analysis and interpretation of the results. | 4.0 | 4.0 |
| 13 | How were quantitative variables handled in the analyses, and if applicable, what groupings were chosen and why?<br>The study has the following aim: to examine the relationship between the percentage of population fully vaccinated and the incidence of new COVID-19 cases in a given population, and to assess the impact of vaccination on the incidence of COVID-19 cases. | To fulfill this aim, the researchers would need to handle the quantitative variable of percentage of population fully vaccinated in their analyses. This could be done in a variety of ways, such as by using statistical tests to determine the strength of the relationship between vaccination and COVID-19 incidence, or by creating graphs or plots to visualize the data.<br>If applicable, the researchers may also choose to group the data into different categories based on the percentage of population fully vaccinated. For example, they may create separate groups for populations with low, medium, and high vaccination rates. The researchers would need to explain the reasoning behind any groupings that they use and how they chose these groupings. This would help to ensure that the analysis is clear and transparent, and that readers can understand the decisions that were made. | 2.3 | 2.7 |
| 14 | What statistical methods should be described, including those used to control for confounding, according to the STROBE guidelines?<br>The study has the following aim: to examine the relationship between the percentage of population fully vaccinated and the incidence of new COVID-19 cases in a given population, and to assess the impact of vaccination on the incidence of COVID-19 cases. | According to the STROBE guidelines, you should describe all statistical methods that you used in your study to examine the relationship between vaccination and COVID-19 incidence, including any methods used to control for confounding factors. This may include methods such as multivariate regression analysis or stratified analysis, depending on the specific design and analysis of your study. It is important to provide a clear and detailed description of your statistical methods in order to allow readers to understand and evaluate the validity and reliability of your findings. | 2.3 | 1.7 |
| 15 | What methods were used to examine subgroups and interactions in the study?<br>The study has the following aim: to examine the relationship between the percentage of population fully vaccinated and the incidence of new COVID-19 cases in a given population, and to assess the impact of vaccination on the incidence of COVID-19 cases. | It is not clear from the information provided how subgroups and interactions were examined in the study. To accurately answer this question, more information about the methods used in the study would be needed. In general, subgroups can be defined by characteristics such as age, gender, or preexisting health conditions, and examining the relationship between the percentage of population fully vaccinated and the incidence of COVID-19 cases in these subgroups can provide insight into how vaccination may differentially impact different groups. Interactions refer to the combined effect of two or more factors on an outcome, and examining interactions can help determine whether the relationship between vaccination and COVID-19 incidence is modified by other factors. Methods that may be used to examine subgroups and interactions in a study include stratified analyses, multivariate regression, and subgroup analyses. | 2.3 | 3.0 |
| 16 | How should missing data be addressed in the report of an observational study according to the STROBE guidelines?<br>The study has the following aim: to examine the relationship between the percentage of population fully vaccinated and the incidence of new COVID-19 cases in a given population, and to assess the impact of vaccination on the incidence of COVID-19 cases. | According to the STROBE guidelines, the report of an observational study should include information about how missing data were addressed in order to ensure the validity and reliability of the results. This might include:<br>Description of missing data: Describe the extent of missing data in the study, including the number and percentage of participants with missing data for each variable of interest.<br>Handling of missing data: Explain how missing data were handled in the analysis, including any methods used to impute or estimate missing values (e.g., multiple imputation, last observation carried forward).<br>Sensitivity analyses: Conduct and report sensitivity analyses to assess the robustness of the results to different assumptions about missing data. This might include comparing the results of the main analysis to those obtained using different imputation methods or excluding participants with missing data.<br>It is also important to clearly state the aim of the study, as you have done, in order to provide context for the analysis and interpretation of the results. | 3.3 | 3.3 |
| 17 | How was loss to follow-up addressed in a cohort study, and how was matching of cases and controls addressed in a case-control study? In a cross-sectional study, how were analytical methods taking into account the sampling strategy described?<br>The study has the following aim: to examine the relationship between the percentage of population fully vaccinated and the incidence of new COVID-19 cases in a given population, and to assess the impact of vaccination on the incidence of COVID-19 cases. | It's important to note that the study you described aims to examine the relationship between the percentage of population fully vaccinated and the incidence of new COVID-19 cases in a given population, and to assess the impact of vaccination on the incidence of COVID-19 cases. The recommendations from the STROBE guidelines that you provided earlier pertain to reporting the methods used in observational studies in general, and do not specifically apply to the study you described.<br>In order to address loss to follow-up in a cohort study, the researchers could have implemented strategies such as regularly contacting participants to check in on their status and ensure that they are still being followed, or including additional measures such as self-reported outcomes to supplement missing data.<br>In a case-control study, matching of cases and controls refers to the process of selecting controls that are similar to the cases in terms of certain characteristics, such as age, sex, or other potential confounders. The researchers could have used a variety of methods to ensure that the cases and controls are well-matched, such as using a 1:1 ratio of cases to controls or using a propensity score matching approach.<br>In a cross-sectional study, the sampling strategy refers to the method used to select the study participants from the target population. The researchers could have used a variety of methods to select the study participants, such as random sampling or convenience sampling. In order to take into account the sampling strategy in the analytical methods, the researchers could have used statistical techniques such as weighting to account for any potential biases introduced by the sampling method. | 3.7 | 2.3 |
| 18 | What should be included in the report of an observational study in order to describe sensitivity analyses according to the STROBE guidelines?<br>The study has the following aim: to examine the relationship between the percentage of population fully vaccinated and the incidence of new COVID-19 cases in a given population, and to assess the impact of vaccination on the incidence of COVID-19 cases. | In order to describe sensitivity analyses in the report of an observational study that aims to examine the relationship between vaccination and COVID-19 cases, you should include the following information according to the STROBE guidelines:<br>Description of sensitivity analyses: Clearly explain the purpose and methods of the sensitivity analyses, including the assumptions and scenarios that were tested.<br>Results of sensitivity analyses: Report the results of the sensitivity analyses, including any changes in the estimates or conclusions of the main analysis.<br>Interpretation of sensitivity analyses: Discuss the implications of the sensitivity analyses for the overall conclusions of the study, including any limitations or uncertainties that they highlight.<br>It is also important to clearly state the aim of the study, as you have done, in order to provide context for the analysis and interpretation of the results. | 2.3 | 2.0 |
| **Results** |  |  |  |  |
| 19 | How can the numbers of individuals at each stage of a study be reported according to the STROBE guidelines, including those potentially eligible, examined for eligibility, confirmed eligible, included in the study, completing follow-up, and analyzed?<br>The study has the following aim: to examine the relationship between the percentage of population fully vaccinated and the incidence of new COVID-19 cases in a given population, and to assess the impact of vaccination on the incidence of COVID-19 cases. | To report the numbers of individuals at each stage of the study according to the STROBE guidelines, you will need to provide information on the following:<br>The number of individuals who were potentially eligible for the study. This includes all individuals who met the inclusion criteria for the study and were considered as candidates for inclusion.<br>The number of individuals who were examined for eligibility. This includes all individuals who were screened to determine if they met the inclusion criteria for the study.<br>The number of individuals who were confirmed eligible for the study. This includes all individuals who were found to meet the inclusion criteria for the study and were accepted for inclusion.<br>The number of individuals who were included in the study. This includes all individuals who were enrolled in the study and participated in the data collection process.<br>The number of individuals who completed follow-up. This includes all individuals who completed the required follow-up period and provided data for the study.<br>The number of individuals who were analyzed in the study. This includes all individuals whose data was used in the analysis and interpretation of the study results.<br>It is important to report these numbers accurately and transparently in order to allow readers to assess the representativeness and generalizability of the study findings. This is particularly important in observational studies, where the sample may not be representative of the entire population due to selection bias. | 3.3 | 3.0 |
| 20 | What are the reasons for non-participation at each stage, according to the STROBE guidelines?<br>The study has the following aim: to examine the relationship between the percentage of population fully vaccinated and the incidence of new COVID-19 cases in a given population, and to assess the impact of vaccination on the incidence of COVID-19 cases. | According to the STROBE guidelines, you should give reasons for non-participation at each stage of your study. This could include reasons such as the individual declining to participate, being ineligible for the study, or being unable to be contacted. In the context of your study examining the relationship between vaccination and COVID-19 cases, specific reasons for non-participation could include individuals who are unable to be vaccinated due to medical contraindications, or those who refuse to be vaccinated for personal or philosophical reasons. It is important to report these reasons in your study in order to provide a complete and transparent description of your study sample and any potential biases that may have affected your results. | 4.3 | 4.0 |
| 21 | What is the purpose of using a flow diagram in an observational study, according to the STROBE guidelines?<br>The study has the following aim: to examine the relationship between the percentage of population fully vaccinated and the incidence of new COVID-19 cases in a given population, and to assess the impact of vaccination on the incidence of COVID-19 cases. | According to the STROBE guidelines, you should consider using a flow diagram in your observational study to provide a clear and detailed overview of the study design and the flow of participants through the various stages of the study. A flow diagram can help to clearly illustrate the recruitment process, the inclusion and exclusion criteria, and the number of participants at each stage of the study. This can help to provide a transparent and complete description of your study sample and the characteristics of the participants included in your analysis. In the context of your study examining the relationship between vaccination and COVID-19 cases, a flow diagram could be particularly useful in helping to clearly illustrate the recruitment process and the number of individuals who were eligible and agreed to participate in the study. | 4.3 | 4.0 |
| 22 | What characteristics of study participants (such as demographic, clinical, and social) and information on exposures and potential confounders should be included in a study according to the STROBE guidelines?<br>The study has the following aim: to examine the relationship between the percentage of population fully vaccinated and the incidence of new COVID-19 cases in a given population, and to assess the impact of vaccination on the incidence of COVID-19 cases. | According to the STROBE guidelines, you should include the following characteristics of study participants in your report:<br>Demographic characteristics: age, sex, race, ethnicity, socio-economic status, etc.<br>Clinical characteristics: pre existing medical conditions, previous vaccination status, etc.<br>Social characteristics: occupation, lifestyle, behaviors that may affect COVID-19 risk (such as smoking, alcohol consumption, etc.), etc.<br>You should also include information on the exposures being studied (in this case, the percentage of population fully vaccinated) and any potential confounders that may affect the relationship between the exposure and the outcome (in this case, the incidence of COVID-19 cases). Potential confounders could include other factors that may be associated with both the exposure and the outcome, such as access to healthcare, population density, and other demographic, clinical, or social characteristics. | 4.0 | 3.7 |
| 23 | According to the STROBE guidelines, how should the number of participants with missing data for each variable of interest be indicated in a report?<br>The study has the following aim: to examine the relationship between the percentage of population fully vaccinated and the incidence of new COVID-19 cases in a given population, and to assess the impact of vaccination on the incidence of COVID-19 cases. | According to the STROBE guidelines, you should report the number of participants with missing data for each variable of interest in your study. This is important because missing data can affect the validity and reliability of your results. You should also describe how missing data were handled in your analysis. For example, you might have used imputation techniques to estimate missing values, or you might have excluded participants with missing data from the analysis.<br>In your study, you should indicate the number of participants with missing data for variables such as vaccination status, COVID-19 status, and any other variables that are relevant to your research question. This will allow readers to understand the potential impact of missing data on your results, and to assess the robustness of your findings. | 5.0 | 4.3 |
| 24 | According to the STROBE guidelines, how should the follow-up time be summarized in a cohort study, including the average and total amount of follow-up?<br>The study has the following aim: to examine the relationship between the percentage of population fully vaccinated and the incidence of new COVID-19 cases in a given population, and to assess the impact of vaccination on the incidence of COVID-19 cases. | In a cohort study, it is important to report the follow-up time to allow readers to understand the duration of the study and the potential for changes in the variables of interest over time. According to the STROBE guidelines, you should summarize the follow-up time in your study by reporting the average and total amount of follow-up for the cohort.<br>For example, you might report the average follow-up time as the total follow-up time divided by the number of participants in the cohort, and the total follow-up time as the length of time from the start of the study to the end of follow-up for the last participant. This will help readers to understand the duration of the study and to assess the potential impact of changes in the variables of interest over time on your results. | 3.7 | 3.3 |
| 25 | According to the STROBE guidelines, how should the numbers of outcome events or summary measures be reported over time in a cohort study?<br>The study has the following aim: to examine the relationship between the percentage of population fully vaccinated and the incidence of new COVID-19 cases in a given population, and to assess the impact of vaccination on the incidence of COVID-19 cases. | According to the STROBE guidelines, in a cohort study, you should report the numbers of outcome events or summary measures over time to allow readers to understand the progression of the outcome of interest in the study population. This is particularly important if the outcome is changing over time, as it allows readers to understand the trajectory of the outcome and to assess the potential impact of the exposure on the outcome.<br>In your study, you should report the numbers of outcome events (such as the incidence of new COVID-19 cases) or summary measures (such as the mean or median number of cases) over time for the vaccinated and unvaccinated groups. This will allow readers to compare the incidence of COVID-19 cases in the two groups and to assess the impact of vaccination on the incidence of COVID-19 cases. You should also report any relevant summary measures of the outcome over time, such as the mean or median number of cases per time period. | 2.7 | 3.3 |
| 26 | According to the STROBE guidelines, how should the numbers in each exposure category or summary measures of exposure be reported in a case-control study?<br>The study has the following aim: to examine the relationship between the percentage of population fully vaccinated and the incidence of new COVID-19 cases in a given population, and to assess the impact of vaccination on the incidence of COVID-19 cases. | According to the STROBE guidelines, in a case-control study, you should report the numbers of participants in each exposure category, or summary measures of exposure, to allow readers to understand the distribution of the exposure of interest in the study population. This is important because the distribution of the exposure may affect the strength and direction of the relationship between the exposure and the outcome.<br>In your study, you should report the numbers of participants in each exposure category (such as fully vaccinated and not fully vaccinated) or summary measures of exposure (such as the mean or median percentage of population fully vaccinated) for the case and control groups. This will allow readers to compare the distribution of the exposure between the two groups and to assess the potential impact of the exposure on the incidence of COVID-19 cases. You should also report any relevant summary measures of the exposure, such as the mean or median percentage of population fully vaccinated in each group. | 4.0 | 3.7 |
| 27 | According to the STROBE guidelines, how should the numbers of outcome events or summary measures be reported in a cross-sectional study?<br>The study has the following aim: to examine the relationship between the percentage of population fully vaccinated and the incidence of new COVID-19 cases in a given population, and to assess the impact of vaccination on the incidence of COVID-19 cases. | According to the STROBE guidelines, in a cross-sectional study, you should report the numbers of outcome events or summary measures to allow readers to understand the prevalence of the outcome of interest in the study population. This is important because the prevalence of the outcome may affect the strength and direction of the relationship between the exposure and the outcome.<br>In your study, you should report the numbers of outcome events (such as the incidence of new COVID-19 cases) or summary measures (such as the mean or median number of cases) for the fully vaccinated and not fully vaccinated groups. This will allow readers to compare the prevalence of the outcome between the two groups and to assess the potential impact of the exposure on the incidence of COVID-19 cases. You should also report any relevant summary measures of the outcome, such as the mean or median number of cases in each group. | 3.7 | 4.0 |
| 28 | According to the STROBE guidelines, how should unadjusted estimates and, if applicable, confounder-adjusted estimates and their precision (such as 95% confidence intervals) be reported, and how should it be made clear which confounders were adjusted for and why they were included?<br>The study has the following aim: to examine the relationship between the percentage of population fully vaccinated and the incidence of new COVID-19 cases in a given population, and to assess the impact of vaccination on the incidence of COVID-19 cases. | According to the STROBE guidelines, you should report unadjusted estimates and, if applicable, confounder-adjusted estimates and their precision (such as 95% confidence intervals) to allow readers to understand the relationship between the exposure and the outcome, and to assess the potential impact of confounders on this relationship.<br>In your study, you should report unadjusted estimates of the relationship between the percentage of population fully vaccinated and the incidence of COVID-19 cases, as well as any confounder-adjusted estimates and their precision. You should also make it clear which confounders were adjusted for in the analysis and why they were included. This will allow readers to understand the impact of the confounders on the relationship between the exposure and the outcome, and to assess the robustness of your findings.<br>For example, you might report the unadjusted odds ratio for the relationship between vaccination and COVID-19 incidence, as well as the confounder-adjusted odds ratio and its 95% confidence interval. You should also specify which confounders were adjusted for in the analysis (such as age, sex, and pre existing medical conditions) and why these confounders were included (such as because they are known to be associated with both the exposure and the outcome). | 4.3 | 3.7 |
| 29 | What are the guidelines for reporting category boundaries when continuous variables are categorized in observational studies in epidemiology?<br>The study has the following aim: to examine the relationship between the percentage of population fully vaccinated and the incidence of new COVID-19 cases in a given population, and to assess the impact of vaccination on the incidence of COVID-19 cases. | According to the STROBE guidelines, when continuous variables are categorized in observational studies, the category boundaries should be reported. In your study examining the relationship between vaccination and COVID-19 incidence, you should report the category boundaries for the percentage of population fully vaccinated. This will allow readers to understand how the continuous variable of vaccination was divided into categories for analysis. | 3.3 | 3.3 |
| 30 | What is the STROBE recommendation for translating estimates of relative risk into absolute risk for a meaningful time period, if relevant?<br>The study has the following aim: to examine the relationship between the percentage of population fully vaccinated and the incidence of new COVID-19 cases in a given population, and to assess the impact of vaccination on the incidence of COVID-19 cases. | According to the STROBE guidelines, if relevant, consider translating estimates of relative risk into absolute risk for a meaningful time period. In your study examining the relationship between vaccination and COVID-19 incidence, this may be relevant if you want to express the risk of developing COVID-19 in terms of the number of cases that occur over a specific time period (e.g. cases per 100,000 population per year). Translating relative risk into absolute risk can help readers better understand the practical implications of the study findings. | 3.3 | 2.3 |
| 31 | What are the guidelines for reporting other analyses done, such as analyses of subgroups and interactions and sensitivity analyses, according to the STROBE guidelines?<br>The study has the following aim: to examine the relationship between the percentage of population fully vaccinated and the incidence of new COVID-19 cases in a given population, and to assess the impact of vaccination on the incidence of COVID-19 cases. | According to the STROBE guidelines, you should report other analyses that were conducted in your study, such as analyses of subgroups (e.g. age, sex, ethnicity) and interactions (e.g. the combined effect of two or more variables) and sensitivity analyses (analyses that test the robustness of the results by altering assumptions or methods). This will allow readers to understand the full range of analyses that were conducted and the potential impact of any factors that were considered. In your study examining the relationship between vaccination and COVID-19 incidence, it may be useful to report any subgroup or interaction analyses that were done to better understand how the relationship between vaccination and COVID-19 incidence varies across different subpopulations or under different conditions. Sensitivity analyses can also be helpful in assessing the robustness of the study findings. | 3.3 | 2.0 |
| **Discussion** |  |  |  |  |
| 32 | According to the STROBE guidelines, how should key results be summarized with reference to the study objectives?<br>The study has the following aim: to examine the relationship between the percentage of population fully vaccinated and the incidence of new COVID-19 cases in a given population, and to assess the impact of vaccination on the incidence of COVID-19 cases. | According to the STROBE guidelines, you should summarize the key results of your study with reference to the study objectives to allow readers to understand the main findings of the study and how they relate to the research question.<br>In your study, you should summarize the key results of the relationship between the percentage of population fully vaccinated and the incidence of COVID-19 cases, with reference to the study objective of assessing the impact of vaccination on the incidence of COVID-19 cases. You should report the main findings of the study, such as the magnitude and direction of the relationship between the exposure and the outcome, and any relevant measures of precision (such as 95% confidence intervals).<br>For example, you might report the unadjusted and confounder-adjusted odds ratios for the relationship between vaccination and COVID-19 incidence, along with their 95% confidence intervals, and summarize the main findings by stating that vaccination was associated with a reduced risk of COVID-19 incidence. You should also highlight any key limitations or strengths of the study, and discuss how the results contribute to the existing literature on the topic. | 4.3 | 4.0 |
| 33 | According to the STROBE guidelines, how should the limitations of the study be discussed, taking into account sources of potential bias or imprecision, and considering both the direction and magnitude of any potential bias?<br>The study has the following aim: to examine the relationship between the percentage of population fully vaccinated and the incidence of new COVID-19 cases in a given population, and to assess the impact of vaccination on the incidence of COVID-19 cases. | According to the STROBE guidelines, you should discuss the limitations of your study to allow readers to understand the potential sources of bias or imprecision that may affect the validity and reliability of your results. You should consider both the direction and magnitude of any potential bias, as this will help readers to understand the potential impact of the bias on the study conclusions.<br>In your study, you should discuss the limitations that may have affected the relationship between the percentage of population fully vaccinated and the incidence of COVID-19 cases, such as potential sources of bias or imprecision. For example, you might discuss the potential for selection bias if the vaccinated and unvaccinated groups differed in important ways that could have affected the outcome, or the potential for measurement error if the vaccination status or COVID-19 status of the participants was not accurately assessed. You should also consider the direction and magnitude of any potential bias, and discuss how this may have affected the study conclusions. This will allow readers to understand the potential limitations of the study and to assess the robustness of your findings. | 5.0 | 4.7 |
| 34 | According to the STROBE guidelines, how should a cautious overall interpretation of the results be given, considering the study objectives, limitations, multiplicity of analyses, results from similar studies, and other relevant evidence?<br>The study has the following aim: to examine the relationship between the percentage of population fully vaccinated and the incidence of new COVID-19 cases in a given population, and to assess the impact of vaccination on the incidence of COVID-19 cases. | According to the STROBE guidelines, you should give a cautious overall interpretation of the results of your study, considering the study objectives, limitations, multiplicity of analyses, results from similar studies, and other relevant evidence. This will allow readers to understand the main findings of the study and to assess the potential implications of the results in the context of the research question.<br>In your study, you should interpret the results of the relationship between the percentage of population fully vaccinated and the incidence of COVID-19 cases, considering the study objective of assessing the impact of vaccination on the incidence of COVID-19 cases. You should discuss the main findings of the study, such as the magnitude and direction of the relationship between the exposure and the outcome, and any relevant measures of precision (such as 95% confidence intervals). You should also consider the limitations of the study, such as potential sources of bias or imprecision, and discuss how these may have affected the study conclusions. You should also consider the results of similar studies and other relevant evidence, and discuss how the findings of your study fit within the broader context of the research on the topic. This will allow readers to understand the potential implications of the results for the relationship between vaccination and COVID-19 incidence. | 3.3 | 2.7 |
| 35 | According to the STROBE guidelines, how should the generalizability (external validity) of the study results be discussed?<br>The study has the following aim: to examine the relationship between the percentage of population fully vaccinated and the incidence of new COVID-19 cases in a given population, and to assess the impact of vaccination on the incidence of COVID-19 cases. | According to the STROBE guidelines, you should discuss the generalizability (external validity) of the study results to allow readers to understand the extent to which the findings of the study can be applied to other populations or settings.<br>In your study, you should discuss the generalizability of the results of the relationship between the percentage of population fully vaccinated and the incidence of COVID-19 cases to other populations or settings. You should consider the characteristics of the study population, such as the age, sex, and preexisting medical conditions of the participants, and discuss how these characteristics may affect the generalizability of the results. You should also consider the setting of the study, such as the geographic location, and discuss how the local context may affect the generalizability of the results.<br>For example, you might discuss whether the findings of your study are likely to be generalizable to other populations with different age, sex, or preexisting medical condition distributions, or to other settings with different geographic or cultural contexts. This will allow readers to understand the potential limitations of the study in terms of generalizability, and to assess the potential relevance of the results to other populations or settings. | 3.7 | 3.3 |
| **Mean overall score** |  |  | 3.6 | 3.3 |

More in details, item #9 (“*What should be included in the definition of outcomes, exposures, predictors, potential confounders, and effect modifiers in an observational study, and how should diagnostic criteria be provided, if applicable, according to the STROBE guidelines?*”), in the Methods section, received a very satisfactory response in terms of both coherence (5.0/5.0) and relevance (5.0/5.0); therefore, the responses given were rated very positively (“*According to the STROBE guidelines, in order to clearly define all outcomes, exposures, predictors, potential confounders, and effect modifiers in your observational study, you should provide detailed descriptions of each of these components. Outcomes: In this study, the outcome would be the incidence of new COVID-19 cases in a given population. You should specify how you will measure the incidence of COVID-19 cases (e.g. number of new cases per 100,000 population), and provide any relevant diagnostic criteria for COVID-19 (e.g. symptoms, test results). Exposures: The exposure in this study would be the percentage of the population that is fully vaccinated. You should specify how you will measure the percentage of the population that is fully vaccinated (e.g. number of individuals who have received both doses of the vaccine), and provide any relevant information on the specific vaccine(s) used. Predictors: The predictor in this study would be the percentage of the population that is fully vaccinated. [*…]”.

Similarly, the answer to item #10, Methods section, was also rated very positively (“*What should be included in the description of sources of data and methods of assessment for each variable of interest in an observational study, and how should comparability of assessment methods be described if there is more than one group, according to the STROBE guidelines?*”), and received a satisfactory response in terms of both coherence (4.7/5.0) and relevance (4.7/5.0). Here, too, in the opinion of the evaluators, the transformer response was complete and adequate (“*According to the STROBE guidelines, in order to provide a clear and transparent description of the sources of data and methods of assessment for each variable of interest in your observational study, you should include the following information: For each variable of interest (e.g. percentage of population fully vaccinated, incidence of new COVID-19 cases), specify the source of data (e.g. administrative records, survey data). For each variable of interest, describe the method of assessment in detail. For example, if you are using administrative records to measure the percentage of population fully vaccinated, you should describe how the records were collected and how you will extract the relevant information. If you are using survey data to measure the incidence of new COVID-19 cases, you should describe the survey instrument and how you will collect and analyze the data. If there is more than one group in your study (e.g. different populations or time periods), describe the comparability of assessment methods for each variable of interest. For example, if you are comparing the percentage of population fully vaccinated in two different populations, you should describe whether the vaccination programs were similar in the two populations and whether the data sources and methods of assessment were comparable. [*…]”.

In contrast, some responses received particularly negative ratings from domain experts. For example, the response to item/recommendation #14, Methods section, received negative comments in terms of both coherence and relevance (2.3/5.0, and 1.7/5.0, respectively). The question was: “*What statistical methods should be described, including those used to control for confounding, according to the STROBE guidelines?*”. And the answer (“*According to the STROBE guidelines, you should describe all statistical methods that you used in your study to examine the relationship between vaccination and COVID-19 incidence, including any methods used to control for confounding factors. This may include methods such as multivariate regression analysis or stratified analysis, depending on the specific design and analysis of your study. It is important to provide a clear and detailed description of your statistical methods in order to allow readers to understand and evaluate the validity and reliability of your findings*.”) was not deemed adequate either in terms of coherence or relevance.

Similarly, for item #18 (“What should be included in the report of an observational study in order to describe sensitivity analyses according to the STROBE guidelines?”), the answer given was also not considered adequate by the evaluators, giving mean scores of 2.3/5.0 and 2.0/5.0.

## 4. Discussion

In this study, we used an open early-stage AI-based transformer, namely ChatGTP [19,20], to reformulate and answer the STROBE recommendations to support the conduction of an epidemiological study. We then assessed the coherence and relevance of the transformer’s outputs. We used an observational study that analyzes publicly available data to investigate the relationship between the percentage of population fully vaccinated and new COVID-19 cases. It is important to note that we started from the premise that while AI can potentially assist in reproducing a study, the research question, the aim of the study, and all the aspects of originality should remain the sole domain of humans. Therefore, we began our simulation by using a research question and aim of the study devised by a human.

After ChatGPT has answered to all the STROBE recommendations, we asked independent experts to qualitatively evaluate the coherence and relevance of the tranformer’s outputs. In the last few months, these innovative systems have proven to be fast and intuitive and an important support for researchers. However, the coherence, relevance, and even the correctness of their answers is not always clear, and there are currently no studies, to our knowledge, that critically analyze the use of these tools in epidemiological research [21]. The choice of the STROBE checklist and the method of turning its recommendations into questions for ChatGPT represents an attempt to standardize and make our analysis as more reproducible as possible. In fact, by following the STROBE guidelines, researchers can help ensure that their results are accurately and clearly reported, which can help other researchers and policymakers understand and interpret the results of the study. This is particularly important in the field of epidemiology, as the results of epidemiological studies can have important implications for public health policy and practice.

From the results of our analysis, we found that ChatGPT can be a valuable support for researchers, both experienced and inexperienced, in setting up an epidemiological study, particularly observational, following internationally recognized guidelines and standards. The average score attributed by experts to the responses given by the AI-based transformer was above 3.5 over 5, but with a fair amount of variability among responses to individual outputs. This is consistent with suggestions and preliminary analyses in the limited literature available on these tools [9–11,22], which indicate that AI-based transformers are beginning to be used but with some skepticism, partly due to their suboptimal ability to respond adequately, in terms of consistency and relevance, to questions posed by domain expert researchers. Therefore, it seems essential that users of these tools have knowledge about the subject matter and a critical mindset when evaluating their outputs. Blindly accepting the answers of these tools may still be too great a risk to the integrity of science and thus human society.

It is also interesting to note that the researchers rated some ChatGPT outputs very positively and others very negatively. Notably, both the responses rated very positively and those rated very negatively were in the methods part. This suggests that, when properly queried, the transformer may provide more appropriate answers in the methodological and data analysis and reporting domains, which were found to have the highest level of consistency and relevance.

Even in the introduction part, this AI-based transformer can be useful in placing a study in context. As literature has shown [21], these systems can function as true “scientific writers” and can quickly and adequately contribute to the writing of entire paragraphs. However, our study found that the responses to STROBE recommendations related to the introduction/background and discussion/interpretation paragraphs of scientific articles did not receive high scores, indicating that the tool is not yet fully adequate to support (or potentially replace) researchers in this part of scientific articles, particularly in epidemiological studies. A possible explanation for this drop in performance may be related to the training data of ChatGPT. Specifically, the AI-based transformer was not specifically trained on scientific articles. This issue may be partially addressed by specific training or fine-tuning [16,23].

Our analysis showed the ability of AI-based transformers to generate answers and human-like text, which could potentially be used to conduct epidemiological studies or write research articles. This could potentially save time and resources for researchers, as these activities can be time-consuming and labor-intensive.

However, there are also several potential drawbacks to the use of AI-based transformers for scientific research [21,24]. One concern is the risk of bias in the data that is used to train these systems. If the data used to train the AI system is biased, the system may produce biased results or make biased recommendations. This could have serious consequences, particularly in the field of public health and epidemiology where decisions about interventions and policies may be based on the results of research.

Another concern that is often reported in the critical analysis of these systems [19,25], is the potential for AI-based transformers to replace human researchers, potentially leading to job loss and the devaluation of human expertise [10]. However, our assessment suggests a possible shift in the “study-experiment-analysis-results-writing-publication” paradigm. It seems more likely that, with the support of these systems, we will move towards a model where the researcher can focus more on the study and experimental phase then on methodological problems and questions.

Another risk associated with the use of AI in scientific research is the potential for fraudulent manipulation of large-scale scientific publications. An example of this is related to the scientific publisher IOP, which retracted as many as 850 articles in 2022 after a researcher at the University of Cambridge in the United Kingdom discovered that many of them contained nonsensical phrases produced by artificial intelligence programs trying to avoid anti-plagiarism software. These phrases were repeated a staggering number of times in different articles from different groups. When IOP began investigating, the editor found other similarities that suggested the articles came from a so-called “Scientific Paper Mill” a company that produces and sells pseudo-articles for a fee.

The use of AI-based solutions capable of generating text in scientific research also brings with it many legal implications. However, it is beyond the scope of this article to address in detail each and all the possible legal issues that may arise when using AI tools to generate text to be published in scientific studies. For our purposes, we can highlight that one of the main concerns is the issue of plagiarism and copyright infringement. AI-generated text may be similar or identical to existing copyrighted material, which could lead to legal issues for researchers and their institutions. It is important for researchers to be aware of these potential issues and take steps to ensure that their work does not violate any copyright laws. Using anti-plagiarism software to evaluate the generated text can be a helpful tool in reducing the risk of publishing non-original text.

We look forward to the development and refinement of these systems, which will be helpful in preserving scientific integrity and ethics and will be of great assistance to publishers and other stakeholders. These software programs are designed to detect similarities between a given text and existing material. They can identify potential instances of plagiarism or copyright infringement, allowing researchers to make necessary changes before publishing their work. It is important to note, however, that anti-plagiarism software may not catch all instances of plagiarism. Therefore, it is important for researchers to also use their own judgment and to carefully review the AI-generated text for any potential instances of plagiarism or copyright infringement.

Another legal implication in using AI-based solutions for text generation in scientific research is the accuracy of the information. As mentioned earlier, AI-generated text may contain errors or inaccuracies that can have serious consequences for the scientific community. For example, if an AI-generated article is published in a reputable journal and it contains inaccurate information, it could lead to other researchers basing their work on flawed data. Therefore, it is crucial for researchers to thoroughly check that the final product is sound and reliable.

In short, it is important to stress that even if AI tools are used to generate text, the responsibility for the research still lies with the humans to whom the work is attributed. This means that researchers and their institutions are responsible for ensuring that their work is legally compliant and that it does not infringe on any copyright laws or contain any inaccuracies. This highlights the need for careful consideration and adherence to ethical and legal guidelines when utilizing AI in scientific research.

### 4.1. Limits of the study

It is important to note that, being an ML algorithm, ChatGPT’s answers may differ if re-run after some time. This represents a limitation in the reproducibility of the results of our study. Furthermore, there is no single way to formulate questions to the transformer; different questions, even on the same topic, produce different answers, which may be more or less consistent and relevant to the objective of the analysis, representing a limitation of the study. However, in the context of the present study, we tried to standardize this process as much as possible by starting with STROBE recommendations and directly asking the AI-based transformer to adapt them into prompts for itself. This may have at least partially mitigated the risk of variability.

## 5. Conclusions

In this exploratory study, we evaluated the coherence and relevance of an open AI-based transformer’s answers regarding questions extracted and reformulated from the STROBE recommendations for observational studies.

From our assessment, the transformer can be considered as a valuable support for researchers, both experienced and inexperienced, in setting up an epidemiological study, following internationally recognized guidelines and standards. The average score attributed by experts to the responses given by the AI-based transformer was high, but with a fair amount of variability among responses to individual outputs.

Responses related to the methods, data analysis and reporting domains and recommendations were found to have the highest level of coherence and relevance, emphasizing that these innovative tools may be primarily useful for methodological and statistical support, rather than for building the conceptual frameworks of the study and the narrative structure of the paper.

Therefore, it seems essential that users of these tools have knowledge about the subject matter and a critical mindset when evaluating their outputs. Blindly accepting the answers of these tools may pose a serious threat to the ethics and integrity of scientific research, including epidemiology. Accordingly, we advocate the necessity for taking a proactive stance towards this evolving context, in order to ensure a careful governance of this inevitable process. Maintaining rigor and reliability of scientific research implies establishing a shared and as much as possible standardized method for conducting studies, analyzing data, and reporting its results. The potential benefits of AI in scientific research are undeniable, but it is crucial that we address the risks, and the ethical and legal consequences associated with its use. This requires a collaborative effort between researchers, policymakers, and other stakeholders to define guidelines and best practices - and monitor their fulfillment - for the ethically and legally correct use of AI in scientific research.

## Data Availability

All data produced in the present study are available upon reasonable request to the authors

## Declarations

## Acknowledgment

This article was human-made. No AI was involved in its conceptualization, writing, or reporting. AI was involved where declared in the methods section.

## Ethics approval

Not needed.

## Consent to participate

Not needed.

## Consent for publication

Not needed.

## Data availability

The datasets generated during and/or analyzed during the current study are available from the corresponding author on reasonable request.

## Authorship contribution statement

All authors made full and substantial contributions. DG and FS contributed to conceptualization, writing, reviewing, and editing; AB, and FT, contributed to data processing, writing, reviewing, and editing; NN contributed to data processing, reviewing and editing; AN and GC contributed to reviewing and editing. All authors have given final approval of the manuscript.

